# Switch-like methylation of functional pathways distinguishes COPD and idiopathic pulmonary fibrosis

**DOI:** 10.64898/2025.12.18.25342312

**Authors:** Katherine H. Shutta, Yichen Huang, Vincent L. Carey, Jeong H. Yun, Brian D. Hobbs, Jack A. Elias, Chun Geun Lee, Kevin K. Brown, Gerard Criner, Kevin Flaherty, Andrew H. Limper, Frank Sciurba, Robert A. Wise, Fernando J. Martinez, Edwin K. Silverman, John Quackenbush, Dawn L. DeMeo

## Abstract

Chronic obstructive pulmonary disease (COPD) and idiopathic pulmonary fibrosis (IPF) are phenotypically divergent disorders arising from similar exposures (including cigarette smoke). Differences in DNA methylation may drive the exposed lung towards COPD vs. IPF. To characterize differential methylation in COPD and IPF lung tissue relative to controls, we conducted epigenome-wide association studies of COPD and IPF in lung tissue from the Lung Tissue Research Consortium (N=1029), adjusting for age, sex, smoke exposure, ancestry, estimated cell type composition, and plate. “Switch probes” were defined as CpGs differentially methylated in COPD vs. control and IPF vs. control in opposite directions. Gaussian graphical models were used to mine network properties of switch probes. Differential methylation of genes related to COPD/IPF in the literature was assessed. Switch probe methylation was compared with previously reported gene expression to identify multi-omic switches. We found 13,313 CpGs were associated with COPD and 43,359 with IPF (3,163 overlapping). We identified 1,091 switch CpGs enriched for endocytosis, glycosphingolipid biosynthesis, and pathways in cancer. 24 genes exhibited multi-omic switch behavior, many related to lipid metabolism (*ACSL1*; *FASN*; *LPCAT1*; *MED27*; *NCOR2*). *LPCAT1* is of particular interest due to its role in maintaining phosphatidylcholine, the majority component of surfactant. Further related to surfactant, we observed strong divergent methylation and expression of *ATP11A*, which facilitates endocytosis of surfactant lipids. CONCLUSIONS Our findings suggest multi-omic switch-like regulation may underlie differential COPD/IPF etiology. Future investigation of *LPCAT1* and *ATP11A* could provide new mechanistic understanding and therapeutic avenues.

## 1. Introduction

Chronic obstructive pulmonary disease (COPD) and idiopathic pulmonary fibrosis (IPF) are phenotypically divergent lung diseases that emerge from similar environmental exposures and with aging. Exposure to cigarette smoke is a primary risk factor for the development of both COPD and IPF[1]; occupational exposures such as dust inhalation have also been implicated in the pathogenesis of both diseases[2].

Both COPD and IPF pose substantial public health burdens in terms of reduced quality of life, years spent living with disability, and premature mortality[3, 4]. Both conditions are also currently irreversible, and treatment primarily involves efforts to slow disease progression and to provide supportive care [5, 6].Targeted molecular therapeutics are a promising avenue for COPD[7] and IPF[8] but a key aspect of making such therapies clinically effective will be a better understanding of the molecular differences driving the divergence of the exposed lung towards COPD or IPF.

Genetic variations have been implicated in COPD and IPF, including variations in *SERPINA1* (COPD), *SFTPA2, SFTPC, MUC5B* (IPF), and telomerase-related genes *TERT* and *TERC* (COPD and IPF). [9–17]. Genome-wide association studies (GWAS) have identified larger sets of genes implicated in COPD [18, 19] and IPF [20, 21]. Four single-nucleotide polymorphisms (SNPs) are of particular interest in that the risk allele for IPF is protective for COPD: rs2076295 and rs2744371 (*DSP*), rs2609255 (*FAM13A*) and rs1981997 (*MAPT-KANSL*) [18, 22].

Despite this accumulating evidence, known genetic variations explain only a small fraction of COPD and IPF cases, and little is known about why certain individuals develop COPD, IPF, or remain spirometrically resilient. A growing body of evidence indicates that DNA methylation plays a critical role in moderating the risk of COPD[23] and IPF[24–26]. However, these investigations have been limited by small sample sizes and often conducted solely in peripheral blood, limiting inference about epigenetic mechanisms proximal to the affected lung tissue.

In this study, we performed an epigenome-wide association study (EWAS) to identify methylation loci associated with COPD and/or IPF in lung tissue. To the best of our knowledge, this work is the largest comparative lung tissue EWAS of IPF and COPD to date. We identified a subset of differentially methylated probes (DMPs) that we refer to as “switch probes”; these DMPs have opposite directions of association with COPD and IPF when compared to controls, suggesting a potential role in divergence towards one disease phenotype or the other. We further identified a subset of switch probes that also exhibit differential gene expression, suggesting multi-omic regulation. Regional analyses, pathway enrichment, and graphical modeling analyses provide further systems-level insights of the divergent etiology of COPD and IPF. Some of the results of these studies have been previously reported in the form of an abstract [27].

## 2. Methods

Full detail of the study methods are available in the Supplement.

### 2.1 Study population

The Lung Tissue Research Consortium (LTRC) is a centralized biobank established by the National Heart, Lung, and Blood Institute (https://biolincc.nhlbi.nih.gov/studies/ltrc/) [28]. From 2005 to 2019, the LTRC enrolled 4,486 subjects and obtained lung tissue samples from 3,333 of these individuals during medically indicated lung resection procedures. The study was approved by institutional review boards at each participating institution, and written informed consent was provided by all subjects. Here, we analyze methylation data generated through the Trans-Omics for Precision Medicine program from 1,063 LTRC participants.

### 2.2 Phenotyping

Control, COPD, and IPF phenotypes were assessed using spirometry, clinical diagnoses, and lung tissue pathology. Controls were defined by normal spirometry (forced expiratory volume in one second (FEV_1_) ≥ 80% predicted and FEV_1_/forced vital capacity (FVC) ≥ 0.7) without additional pulmonary diagnosis (see supplement). COPD cases were defined by abnormal spirometry (FEV_1_ < 80% predicted and FEV_1_/FVC < 0.7) without other pulmonary diagnosis (see supplement). IPF was defined by an expert summative clinical diagnosis, in most cases corresponding to pathology.

### 2.3 Methylation array processing

Preprocessing of methylation array data followed a standard workflow (Supplementary Figure E1). The minfi R package was used for noob-adjustment[29], the rcp algorithm was used to correct for probe type bias[30], and functional normalization was performed using the EnMix R package[31]. Methylation beta values were converted to M values using the base-2 logit function.

### 2.4 EWAS statistical methods

Methylation M-values were regressed against an indicator of control vs. lung disease (COPD or IPF) and covariates using limma [32]. Two separate EWAS were conducted: one of COPD vs. control and one of IPF vs. control. Models were adjusted for age, sex, cigarette smoke exposure, ancestry principal components (PCs), estimated cell type composition, and plate. Cigarette smoke exposure was assessed by cg05575921 (*AHRR*) methylation [33]. Ancestry PCs were estimated from whole-genome sequencing using Locating Ancestry from SEquence Reads (LASER) [34, 35]. Cell type composition was estimated using EpiSCORE [36]. EWAS test statistics were corrected for bias and inflation using BACON[37]. Regional differential methylation analysis was performed using DMRcate[38]. Enrichment analyses were conducted with missMethyl [39] and fgsea[40].

### 2.5 GGM methods

Gaussian graphical models (GGMs) were estimated using the graphical lasso[41] (R package huge[42]). To incorporate covariates, GGMs were fit on the residuals obtained by regressing switch probe methylation M-values against the EWAS model covariates. The igraph package [43] was used for community detection and hub score calculation, and fgsea [40] was used for module enrichment analysis.

### 2.6 Literature integration (including DMGs)

To assess DNA methylation signals among genes that have previously been reported to be implicated in COPD and IPF, we manually curated 15 gene sets consisting of 132 unique genes in total. Differentially methylated genes were defined based on a Fisher’s exact test, accounting for array probe bias.

### 2.7 Multi-omic switch methods

We performed a targeted multi-omic integration of switch probe methylation with previous differential gene expression analyses in LTRC lung tissue[28]. To interrogate the function of differentially expressed genes annotated to the switch probes, we used Enrichr [44] to map to BioPlanet pathways [45].

## 3. Results

### 3.1 Participants

Lung tissue DNA methylation profiles were analyzed for 1029 subjects (338 controls, 460 cases with COPD, 231 cases with IPF) (Figure 1, Table 1). Participants were of similar ages across phenotypes. In both COPD and IPF, most cases were male; most controls were female. We observed higher smoke exposure in COPD than in IPF or controls as assessed by lower AHRR methylation, which was negatively associated with self-reported smoking [33] (Table 1, Supplementary Figure E1).

**Figure 1a:**
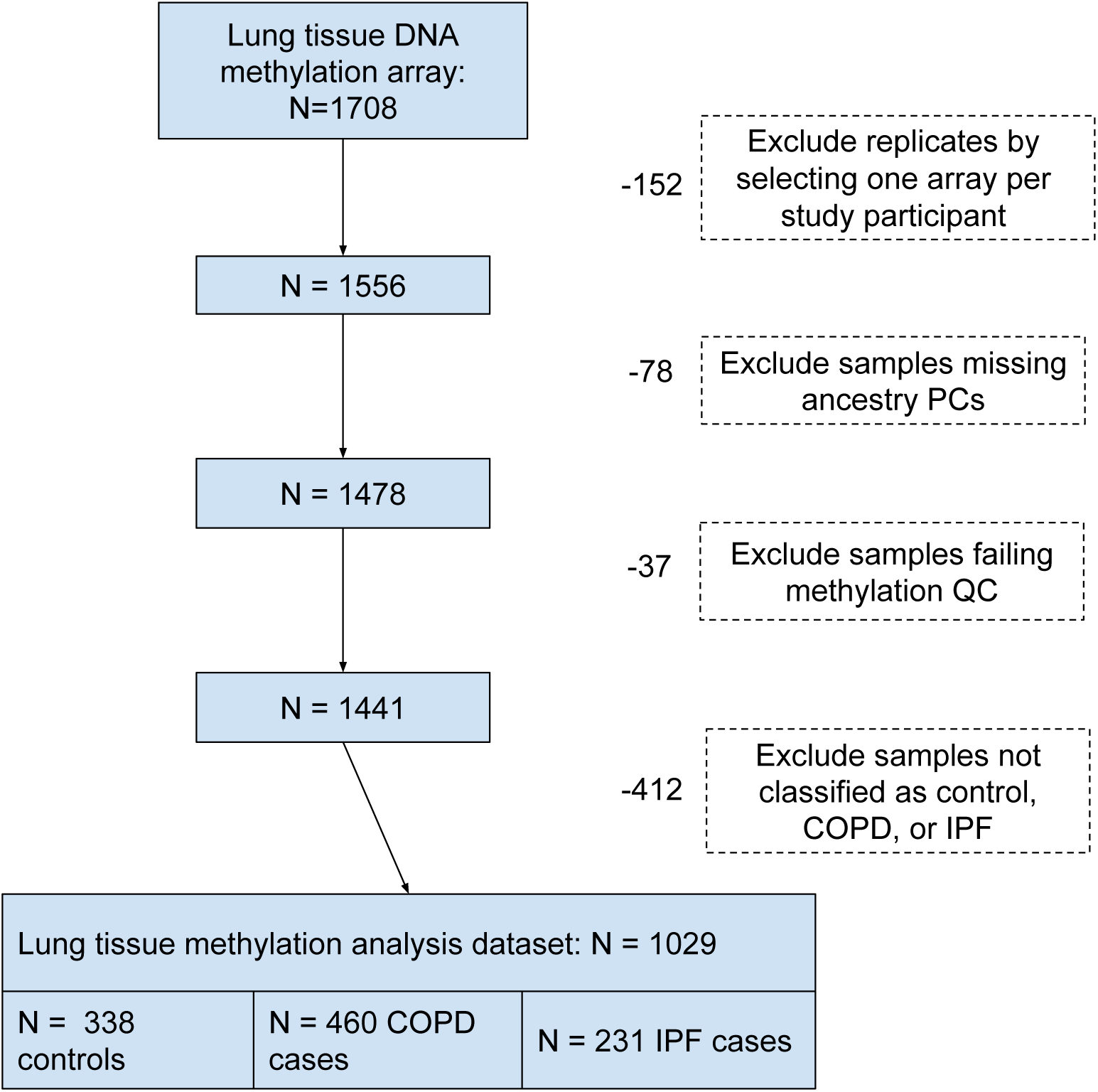
LTRC methylation data sample availability and exclusion criteria.

**Table 1.** Basic characteristics of lung tissue methylation samples, obtained at or near the time of lung resection.

|  | Control<br>(N=338) | COPD<br>(N=460) | IPF<br>(N=231) | Overall<br>(N=1029) |
| --- | --- | --- | --- | --- |
| **Age** |  |  |  |  |
| Mean (SD) | 62.1 (12.1) | 63.1 (9.33) | 63.9 (8.50) | 62.9 (10.2) |
| Median [Min, Max] | 64.0 [24.0, 88.0] | 63.0 [28.0, 85.0] | 64.0 [37.0, 82.0] | 64.0 [24.0, 88.0] |
| **Sex** |  |  |  |  |
| Male | 133 (39.3%) | 254 (55.2%) | 159 (68.8%) | 546 (53.1%) |
| Female | 205 (60.7%) | 206 (44.8%) | 72 (31.2%) | 483 (46.9%) |
| **Race/Ethnicity** |  |  |  |  |
| Non-White | 31 (9.2%) | 42 (9.1%) | 21 (9.1%) | 94 (9.1%) |
| White | 307 (90.8%) | 418 (90.9%) | 210 (90.9%) | 935 (90.9%) |
| **Smoking Status** |  |  |  |  |
| Never | 103 (30.5%) | 21 (4.6%) | 76 (32.9%) | 200 (19.4%) |
| Former | 190 (56.2%) | 378 (82.2%) | 135 (58.4%) | 703 (68.3%) |
| Current | 16 (4.7%) | 32 (7.0%) | 2 (0.9%) | 50 (4.9%) |
| Missing | 29 (8.6%) | 29 (6.3%) | 18 (7.8%) | 76 (7.4%) |
| **AHRR Methylation Beta** |  |  |  |  |
| Mean (SD) | 0.720 (0.0584) | 0.692 (0.0502) | 0.752 (0.0579) | 0.715 (0.0596) |
| Median [Min, Max] | 0.723 [0.527, 0.888] | 0.693 [0.546, 0.840] | 0.751 [0.554, 0.910] | 0.714 [0.527, 0.910] |

### 3.2 Epigenome-wide associations

After quality control measures, 790,856 probes were analyzed for association with COPD and IPF (Figure 1b, Figure 2, Table 2a). The methylation-based epigenetic signature of IPF was generally stronger than that of COPD: 1.7% of probes were significantly differentially methylated in COPD vs. control and 5.5% in IPF vs. control (differentially methylated probes, or DMPs; FDR < 0.05; Figure 2a-b, Supplementary Figure E2a). We observed differences in methylation-estimated cell type composition by disease (Supplementary Figure E2b).

**Figure 1b:**
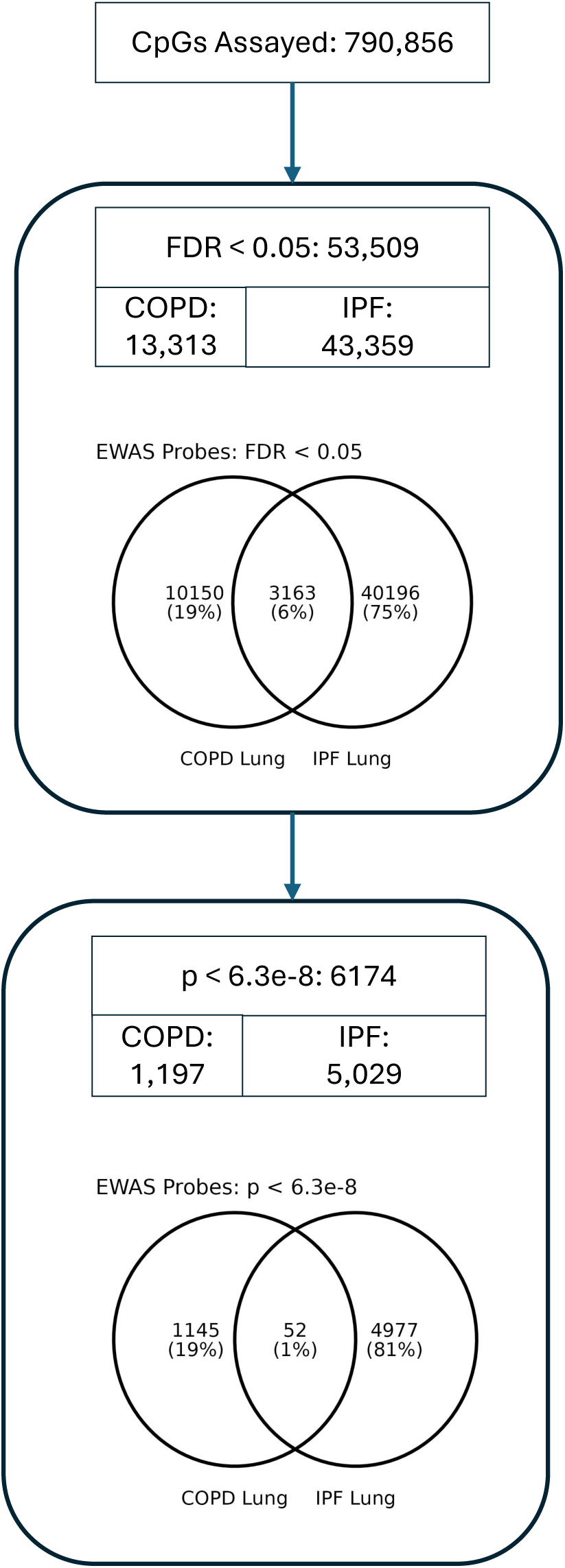
Summary and overlap of EWAS results. Analyses compared COPD vs. control and IPF vs. control samples.

**Figure 2:**
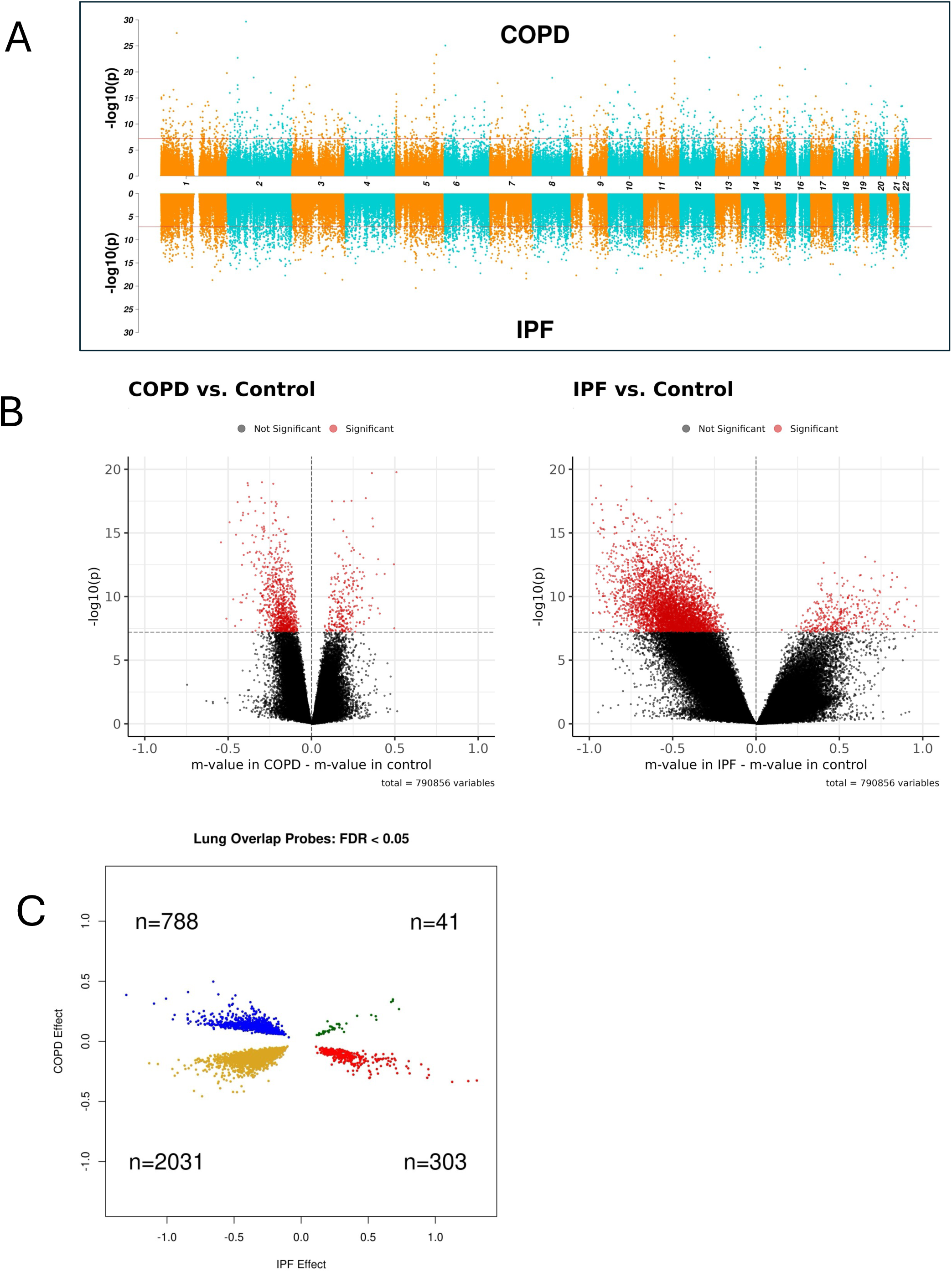
EWAS results. (a) Lung tissue EWAS results in COPD (top) and IPF (bottom)., The red horizontal line indicates the genome-wide significance threshold of p<6.3e-8. (b) Volcano plots of EWAS results in lung tissue. Horizontal line indicates genome-wide significance (c) Effects for EWAS hits significant in both COPD and IPF lung tissue (“overlap probes”). Effects are on the m-value scale.

**Table 2.** (a) Number of hits at FDR < 0.05 and genome-wide significance (p < 6.3e-8) compared to control subjects. Overlap probes are significant in both COPD and IPF. Switch probes are the subset of overlap probes with opposite directions of effect in the two conditions. (b) 10 genome-wide significant switch probes in lung tissue.

|  | Lung -<br>FDR | Lung -<br>Genome<br>Wide |
| --- | --- | --- |
| COPD | 13,313 | 1,197 |
| IPF | 43,359 | 5,029 |
| Overlap | 3,163 | 52 |
| Switch | 1,091 | 10 |

(b)
| probeID | chr | start | genes | COPD<br>effect | COPD p | COPD<br>FDR | IPF effect | IPF p | IPF FDR |
| --- | --- | --- | --- | --- | --- | --- | --- | --- | --- |
| cg02845735 | chr3 | 71,249,393 | AC097634.4;<br>FOXP1 | 0.19 | 6.58e-13 | 3.36e-09 | -0.41 | 1.80e-08 | 3.66e-06 |
| cg06307277 | chr3 | 184,619,647 |  | 0.18 | 3.65e-08 | 2.73e-05 | -0.49 | 5.84e-09 | 1.49e-06 |
| cg10477588 | chr10 | 50,322,279 | SGMS1 | 0.22 | 1.33e-08 | 1.22e-05 | -0.54 | 2.96e-09 | 8.85e-07 |
| cg00452553 | chr11 | 8,779,080 | AC026894.1;<br>DENND2B | 0.16 | 7.50e-10 | 1.15e-06 | -0.36 | 5.87e-09 | 1.50e-06 |
| cg09180057 | chr14 | 89,221,644 | FOXN3 | 0.36 | 1.89e-25 | 2.99e-20 | -0.51 | 2.39e-09 | 7.49e-07 |
| cg24011488 | chr14 | 67,857,957 | RAD51B | 0.22 | 5.45e-10 | 8.83e-07 | -0.67 | 2.52e-10 | 1.39e-07 |
| cg09213451 | chr14 | 61,103,040 |  | 0.20 | 4.60e-08 | 3.25e-05 | -0.52 | 2.06e-09 | 6.69e-07 |
| cg02549846 | chr16 | 12,030,332 | SNX29 | 0.20 | 3.61e-08 | 2.70e-05 | -0.65 | 3.05e-09 | 9.02e-07 |
| cg17003260 | chr17 | 9,004,243 |  | -0.27 | 4.72e-08 | 3.32e-05 | 0.81 | 1.76e-11 | 1.82e-08 |
| cg06647001 | chr20 | 23,097,270 |  | 0.23 | 1.83e-09 | 2.38e-06 | -0.54 | 2.22e-09 | 7.08e-07 |

Of particular interest are DMPs significant in both COPD and IPF (“overlap DMPs”), and the subset of these with opposite direction of effect (“switch DMPs”). Based on the threshold of FDR < 0.05, we identified 1,091 switch DMPs, ten of which also met Bonferroni genome-wide significance (Table 2, Figure 2e, Supplementary Table E1-2).

### 3.3 Regional analysis with DMRcate

Differentially methylated regions (DMRs) were identified using DMRcate (Methods 4.7, Table 3; Supplementary Figure E3; Supplementary Table E3-4). Of these, 354 were “switch DMRs” in which average regional methylation had opposing directions in COPD and IPF vs. control. Protein-coding genes annotated to top switch DMRs include *ATP11A*, *CTBP1, LMNA*, *PIGT*, and *DENND2B*.

**Table 3.** (a) Number of DMRs detected by disease. (b) In lung tissue, top “switch DMRs” (HMFDR < 1e-4 in both COPD and IPF) of the 354 total switch DMRs, which have opposite directions of differential methylation (“mean diff”).(c) KEGG legacy pathways overrepresented at the gene level in the switch DMRs (FDR < 0.05).

| EWAS | #DMR |
| --- | --- |
| COPD lung | 2,384 |
| IPF lung | 8,365 |

(b)
| COPD<br>mean<br>diff | COPD<br>HMFDR | IPF<br>mean<br>diff | IPF<br>HMFDR | Overlapping Genes* |
| --- | --- | --- | --- | --- |
| -0.18 | 1.01e-06 | 0.33 | 1.32e-05 | ATP11A |
| -0.09 | 9.85e-05 | 0.15 | 4.69e-06 | TBCD, AC068014.1 |
| -0.05 | 6.69e-06 | 0.21 | 7.44e-05 | ACSS1 |
| 0.10 | 1.95e-05 | -0.24 | 7.16e-06 | CTBP1-AS, CTBP1 |
| 0.10 | 8.81e-08 | -0.43 | 5.55e-05 |  |
| 0.11 | 5.46e-05 | -0.37 | 4.48e-06 |  |
| 0.11 | 5.66e-06 | -0.21 | 8.20e-06 | C5orf64 |
| 0.13 | 2.49e-07 | -0.28 | 1.88e-05 | LMNA |
| 0.14 | 9.81e-07 | -0.39 | 2.98e-06 |  |
| 0.16 | 4.52e-08 | -0.35 | 3.15e-05 | PIGT |
| 0.17 | 1.13e-06 | -0.31 | 3.00e-06 | AC026894.1, DENND2B |
| 0.17 | 2.82e-06 | -0.35 | 9.89e-07 |  |

\*Overlapping genes were the same for COPD and IPF for all regions shown

(c).
| KEGG gene set | p | FDR | overla<br>p | size | overlapping genes |
| --- | --- | --- | --- | --- | --- |
| hsa04330: Notch signaling pathway | 2.6e-06 | 3.8e-04 | 6 | 47 | CTBP1; DLL1; LFNG; MAML2;<br>NOTCH1; NOTCH4 |
| hsa05200: Pathways in cancer | 4.1e-06 | 3.8e-04 | 13 | 324 | BCL2; CBL; CTBP1; DAPK2; FGF1;<br>FGF14; FGFR2; LAMA5; PLD1;<br>RALGDS; RUNX1; RUNX1T1; TCF7 |
| hsa04910: Insulin signaling pathway | 1.5e-04 | 9.6e-03 | 7 | 136 | CBL; FASN; PRKAG2; PRKAR1B;<br>RAPGEF1; RPTOR; TSC2 |

### 3.4 Over-representation analyses of DMPs and DMRs

We conducted over-representation analysis of DMPs (FDR < 0.05) and of switch DMPs/DMRs (Figure 3a-b, Supplementary Table E5, Supplementary Table E6-7). 107 of 367 candidate KEGG pathways were enriched in IPF and 69 in COPD (44 overlapping at p < 0.05; Figure 3c). Top IPF pathways included hsa04015: Rap1 signaling (p = 2.2e-6), hsa04820: Cytoskeleton in muscle cells (p = 1.1e-5), and hsa04360: Axon guidance (p = 1.2e-5). Top COPD pathways were hsa04015:Rap 1 signaling pathway (p=1.6e-4), hsa04928: Parathyroid hormone synthesis, secretion and action (p = 1.7e-4), and hsa04014: Ras signaling pathway (p = 5.3e-4).

**Figure 3:**
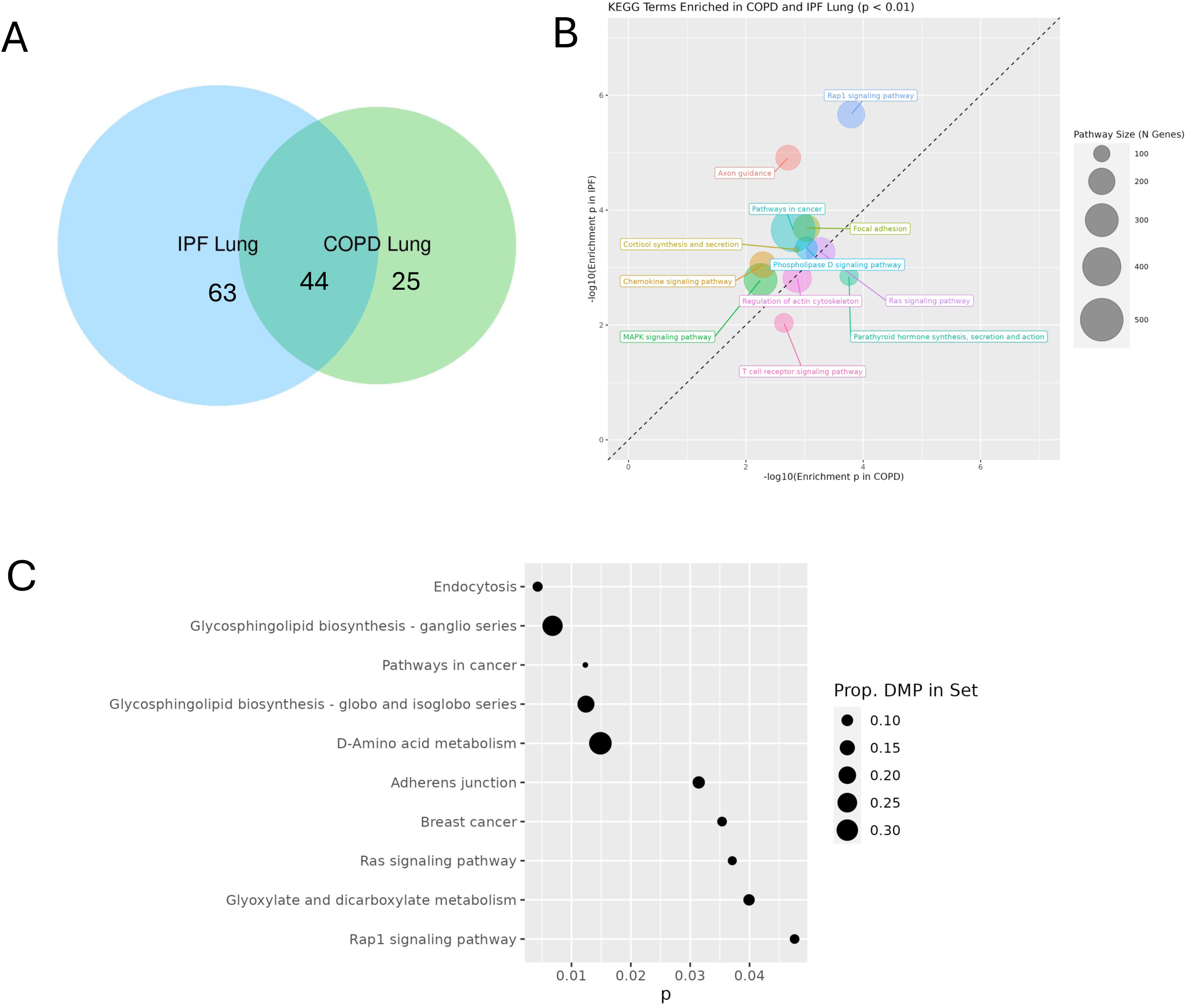
(a) Cross-disease overlap of enrichment (KEGG pathways with p < 0.05) (b) KEGG pathways highly enriched in both COPD and IPF lung (p < 0.01). Bubble size is proportional to KEGG pathway size. (c) Pathways enriched among 1091 lung switch DMPs.

In the 1,091 switch probes, ten KEGG pathways were enriched (p < 0.05; Figure 3e; Supplementary Table E6). The top three pathways were hsa04144: endocytosis (p = 4.3e-3), hsa00604: Glycosphingolipid biosynthesis - ganglio series (p = 6.8e-3), and hsa05200: Pathways in cancer (p = 0.012).

Three of 186 total gene sets were over-represented in the switch DMRs (FDR < 0.05; Table 3c; Supplementary Table E7): hsa04330:Notch signaling pathway (FDR = 3.8e-4), hsa05200: pathways in cancer (FDR = 3.8e-4), and the hsa04910: insulin signaling pathway (FDR = 9.6e-3).

### 3.5 Graphical modeling of switch probes

We constructed Gaussian graphical models (GGMs) using the 1,091 switch probes. (Figure 4, Supplementary Table E8a; Supplementary Figure E4). To understand the role of EWAS hits in the network context, we compared the hub score vs. the −log10(EWAS p-value) for each switch probe (Figure 4c-d). The Spearman correlation between hub score and the −log10(p) was 0.15 (p = 9.8e-7) in COPD and −0.37 (p < 2.2e-16) in IPF, indicating that switch probes tend to serve as network hubs in COPD and are typically less central to the network in IPF. Figure 4c-d highlights four probes of interest: cg03977586 (*WASHC4*), cg07262457, cg17200083 (*PLXNA2*) and cg09180057 (*FOXN3*), discussed in detail in the Supplement (Section 5.1). Community detection identified “switch modules” within the switch probe networks, along with locally influential CpGs (“local hubs”) and within-module KEGG pathway enrichment (Table 4, Figure 4, Methods, Supplementary Table E8b-c). Several switch modules were enriched for the same pathways in both COPD and IPF, including Ras signaling, Rap1 signaling, PI3K-Akt signaling, and pathways in cancer. Local hubs exhibited some disease specificity; for example, *BCL2* is the local hub for the module enriched for Ras/Rap/PI3K-Akt/Pathways in cancer in COPD while *RFLNA* is the local hub for this module in IPF.

**Figure 4:**
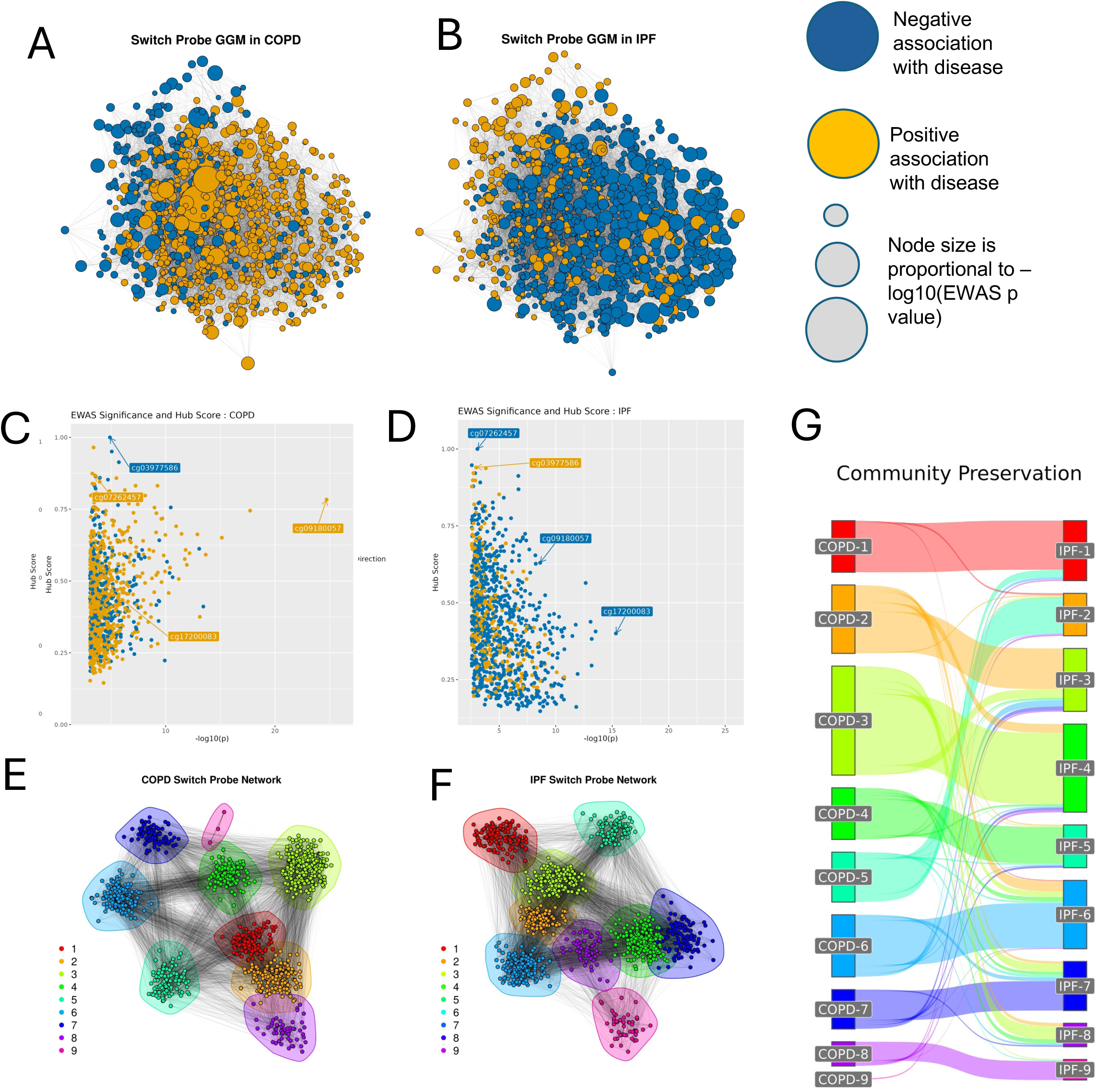
Graphical modeling of switch probes in lung. (a,b) Gaussian graphical models of methylation at the 1082 lung tissue switch probes in COPD (a) and IPF (b). Nodes represent CpG sites, and edges represent partial correlation between methylation at these sites after adjusting for EWAS model covariates (age, sex, cigarette smoke exposure, ancestry, estimated cell type composition, and plate). Node size is proportional to - log(EWAS p-value); larger nodes correspond to more significant EWAS hits and vice-versa. (c-d) Hub score in relation to EWAS significance in COPD (c) and IPF (d). EWAS significance and hub score have a positive Spearman correlation in COPD and a negative correlation in IPF. (e) Communities detected in the COPD switch probe network. (f) Communities detected in the IPF switch probe network. (g) Sankey plot demonstrating preservation and differences between community structures in COPD and IPF.

**Table 4:**
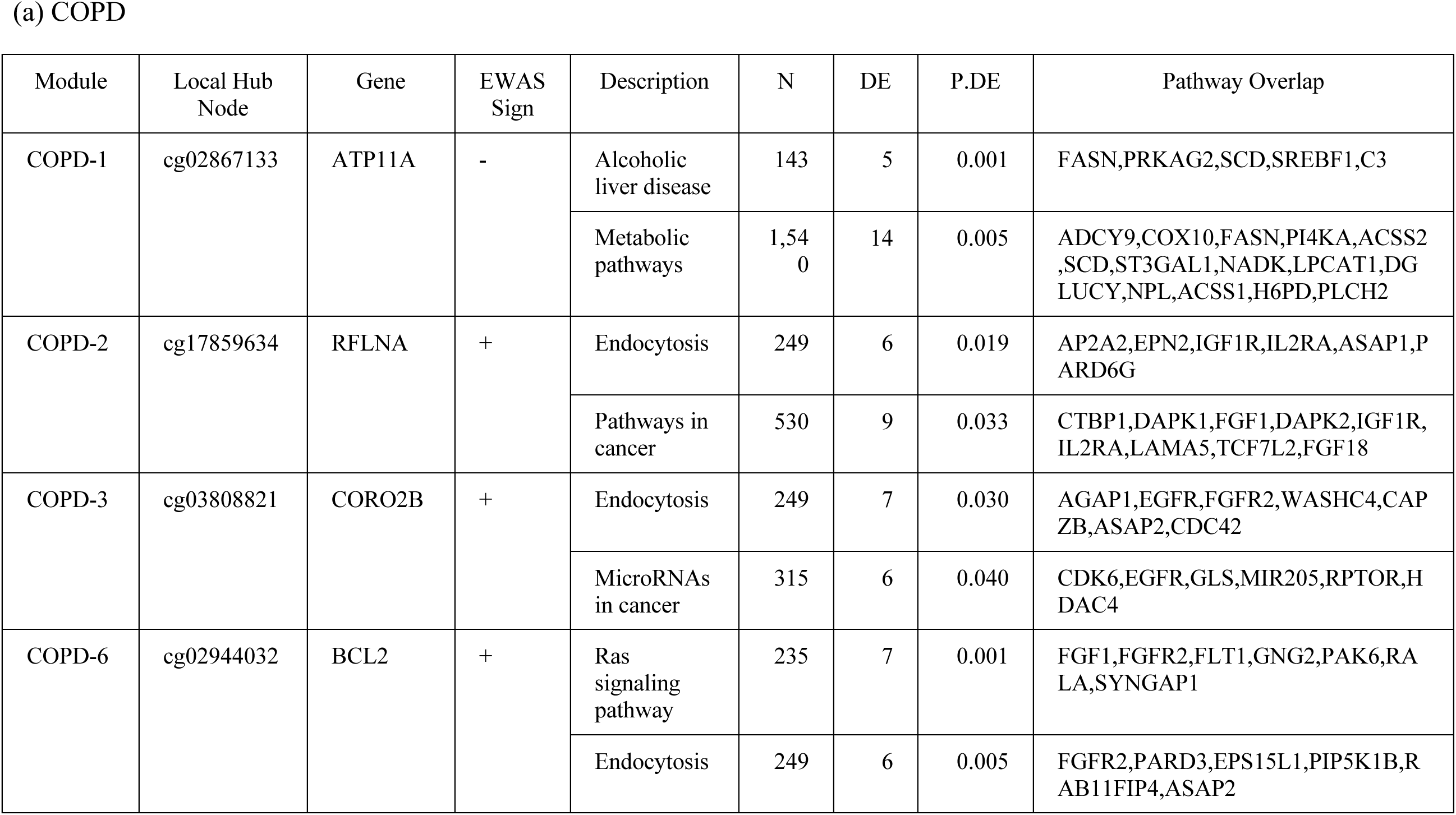
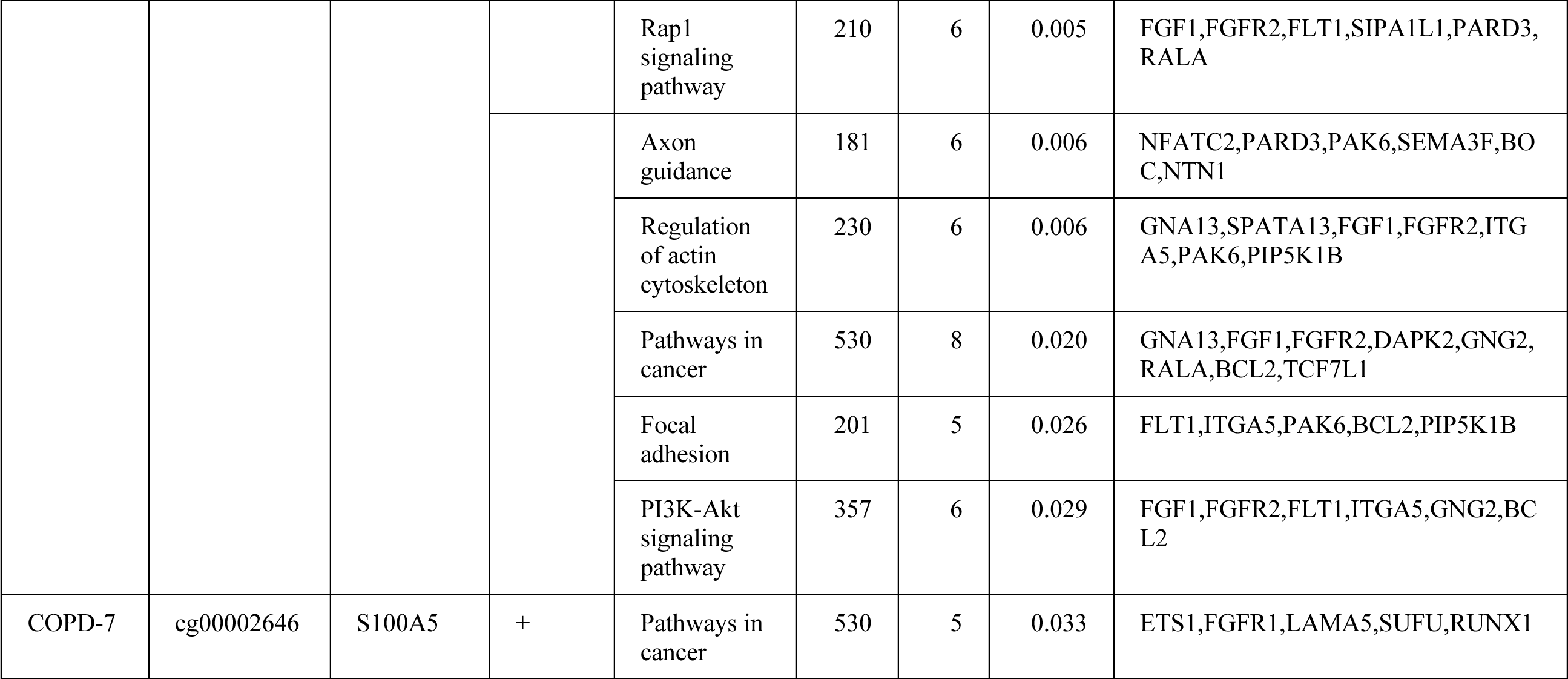
Summary of key switch module characteristics in (a) COPD and (b) IPF. Modules shown are enriched for at least one KEGG pathway at p < 0.05 and with at least five genes overlapping the pathway. Local hub nodes refer to the switch probe with the highest influence in the module according to hub centrality. Module labels match those depicted in Figure 5.

### 3.6 DNA methylation near genes associated with COPD and IPF in previous studies

To assess epigenetic signals among genes previously been reported to be implicated in COPD and IPF, we compiled a list of genes from relevant literature (Methods, Supplementary Table E9). In total, 6,399 probes from the EPIC array annotated to 129 of these “literature genes”. We identified differentially methylated genes (DMGs) based on the significance of the number of DMPs per gene (Methods 4.10). Three literature DMGs met FDR < 0.05 in COPD and seven in IPF, with one overlapping (*ATP11A*;Table 5, Figure 5). Conducting the analysis on all genes with at least one DMP annotated (N=16,694 genes), we found 193 DMGs in COPD and 581 in IPF (Supplementary Section 5.3, Supplementary Table E10).

**Figure 5:**
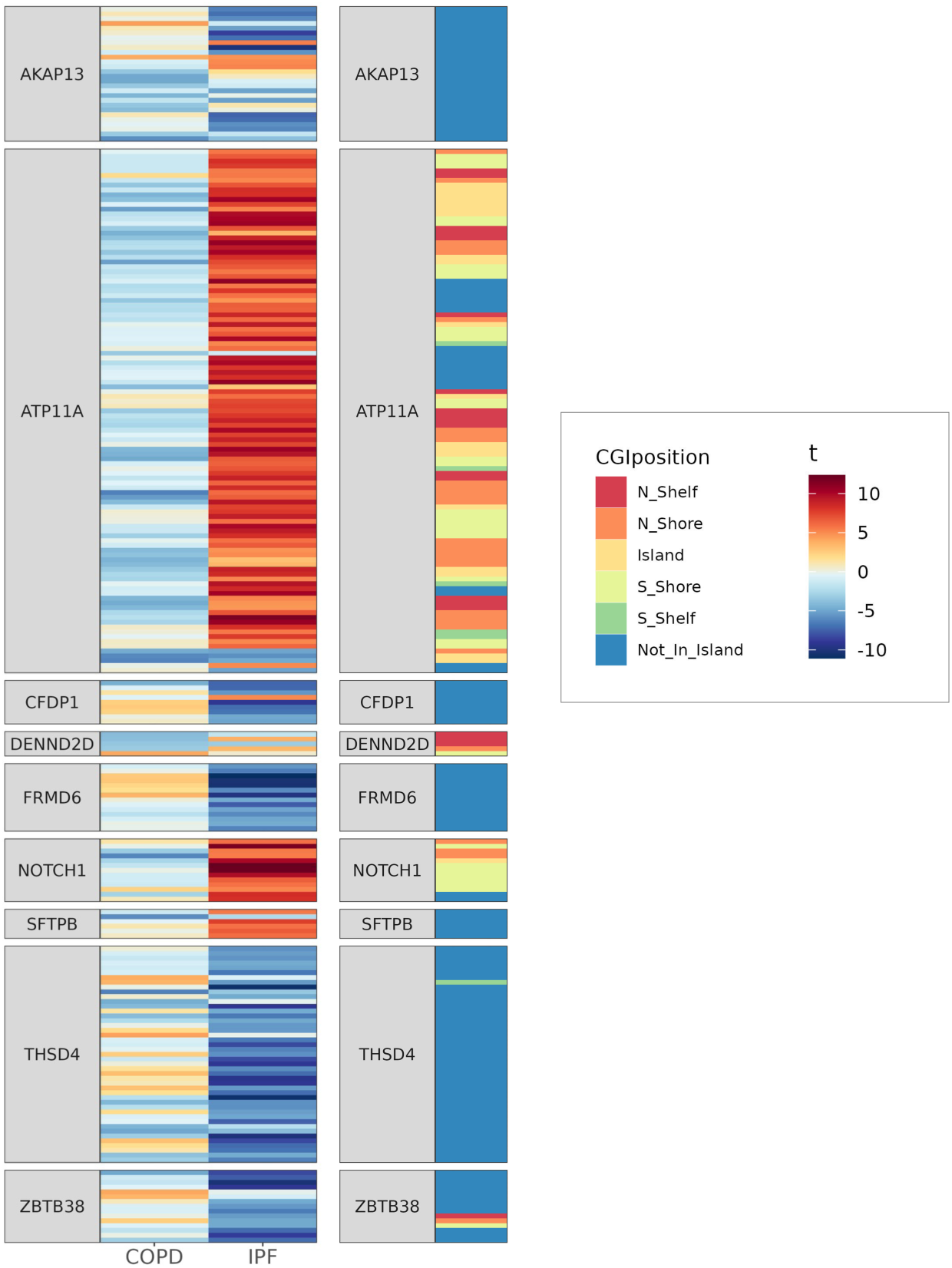
Heatmap of lung tissue DMPs mapped to significantly differentially methylated genes (gene-level Fisher test FDR < 0.05 in either COPD or IPF) that have previously been recognized to be associated with COPD and/or IPF in the literature. Probes shown are significant in lung tissue at least one of the two phenotypes (probe-level t-test FDR < 0.05). Red indicates higher methylation in cases vs. controls while blue indicates lower methylation in cases vs. controls.

### 3.7 Multi-omic switches identified via integration of DNA methylation and gene expression

The lung tissue transcriptomes of COPD and IPF have been previously analyzed in this cohort [Ghosh et al. 2019]. We cross-referenced our EWAS switch probe results with the differential expression results of Ghosh et al. to identify genes exhibiting switch-like behavior between COPD and IPF in both the epigenome and the transcriptome (Methods 4.12; Supplementary Figure E9). The transcriptomic data include 770 of the participants used in our EWAS (240 control, 345 COPD, 185 IPF samples).

184 of our 1,091 switch probes mapped to genes differentially expressed in both in COPD and IPF. Among these 184, we searched for “canonical epigenetic switch probes” (CES probes) that followed the canonical direction of epigenetic regulation via DNA methylation: higher methylation corresponding to lower gene expression and vice-versa. We identified 34 CES probes, which mapped to 24 unique genes, including seven genes with multiple CES probes: 4 mapped to *NCOA7*, 3 mapped to *LPCAT1*, and 2 mapped to *CLSTN1*, *NCOR2*, *NOTCH1*, *RHOBTB2*, and *TNRC18* (Supplementary Figure E9-10; Supplementary Table E12-13). This cross-disease signal provides multi-omic evidence that these 34 CpG sites (24 genes) function as switches in the etiology of COPD and IPF. Mapping these genes to BioPathways gene sets yielded overlap with fatty acid, triacylglycerol, and ketone body metabolism (*NCOR2*; *ACSL1*; *FASN*; *LPCAT1*; *MED27*), triglyceride biosynthesis (*ACSL1*; *FASN*; *LPCAT1*), Fatty acyl-CoA biosynthesis (*ACSL1*; *FASN*), lipid and lipoprotein metabolism (*NCOR2*; *ACSL1*; *FASN*; *LPCAT1*; *MED27*), and lipid metabolism regulation by peroxisome proliferator-activated receptor alpha (PPAR-alpha) (*NCOR2*; *ACSL1*; *MED27*).

## 4. Discussion

We conducted what is, to our knowledge, the largest lung tissue epigenome-wide association study of COPD and IPF to date, using samples obtained from the Lung Tissue Research Consortium (LTRC) with diagnoses based on pathology and expert phenotyping reviewed by respiratory physicians. Our study provides an epigenetic perspective as to why individuals with common environmental exposures go on to develop different lung diseases. To this end, we identified a set of 1,091 differentially methylated “switch probes” with statistically significant and opposing directions of association with COPD vs IPF, 34 of which also exhibited switch-like differential gene expression. Top switch probes mapped to genes related to Wnt signaling (*FOXN3* [46]), the epithelial-mesenchymal transition (EMT; *FOXN3* [47]*, FOXP1* [48]), and cancer development and progression (*DENND2B* [49]*, FOXP1* [48], FOXN3 [46]*, RAD51B* [50]*, SNX29* [51]). We also identified 354 switch differentially methylated regions (“switch DMRs”), including top regions overlapping genes related to the EMT (*LMNA* [52]), cancer (*CTBP1* [53]*, DENND2B* [49]*, LMNA* [54]*, PIGT [55])*, and telomere function (*LMNA* [56]). Switch DMRs were enriched for Notch signaling, pathways in cancer, and insulin signaling. Further evidence implicating Notch/Wnt signaling, the EMT, and telomere function in COPD and IPF are discussed in the Supplement. Differentially methylated genes of interest in COPD and IPF based on previous literature in other omics data types include *ATP11A*, *AKAP13*, and *THSD4* (further discussion in the Supplement).

Network methods provide a non-reductive approach to identifying new insights into disease pathogenesis from omics data that exhibit complex, informative correlations[57]. Using Gaussian graphical models (GGMs), we identified network relationships between the 1,091 switch probes. Leveraging network science methods to quantify node influence, we discovered that in COPD, more influential switch probes tend to have higher EWAS significance while in IPF, the opposite relationship holds. These observations support a characterization of COPD as a disease of epigenetic dysfunction in specific molecular pathways and of IPF as a condition with wide-ranging epigenetic effects in the lung; these may have future implication for therapeutic targeting. We hypothesized that the modular structure of the estimated COPD and IPF switch networks could have important roles in the divergence of the lung toward COPD and IPF. We used community detection to identify “switch modules”, several of which were enriched for metabolic processes, pathways in cancer, Ras signaling, Rap1 signaling, and PI3K/Akt signaling in both COPD and IPF. Local hubs in switch modules identified influential probes mapped to genes that could be promising therapeutic targets, including *ATP11A* and *RFLNA* (COPD and IPF), *BCL2, CORO2B,* and *S100A5* (COPD), and *FOXP1* and *HEXB* (IPF).

Across multiple lines of investigation, including DMPs, DMRs, DMGs, and graphical models, *ATP11A* repeatedly achieved high significance and demonstrated switchlike behavior between COPD and IPF. *ATP11A* encodes a flippase involved in cross-membrane lipid transport; known substrates include phosphatidylserine (PS) and phosphatidylethanolamine (PE), both components of pulmonary surfactant [58, 59]. Sixteen of 1,091 switch probes are mapped to *ATP11A* with generally lower methylation in COPD vs. controls and higher methylation in IPF vs. controls. In both COPD and IPF, *ATP11A* was a switch module hub in a module enriched for KEGG pathways related to metabolism. There is also transcriptomic evidence for the involvement of *ATP11A* in both COPD and IPF in LTRC lung tissue RNA-seq data ([28]; additional data obtained from the authors on request). *ATP11A* expression is higher in COPD than in controls (log fold change = 0.054, FDR = 0.025) and lower in IPF than in controls (log fold change = −0.412, FDR < 2.2e-16). These findings are corroborated in COPD by a recent systematic review [23], which noted that differential methylation of *ATP11A* was associated with COPD vs. control in three different studies across multiple respiratory tissues: lung tissue, small airway epithelia, and sputum.

We observed further evidence implicating pulmonary surfactant lipids in the divergence between COPD and IPF in our multi-omic integration, which revealed a robust set of 34 CpG sites implicated in epigenetic regulation of transcription of 24 key switch genes, many of which are related to lipid biosynthesis and metabolism (*NCOR2*; *ACSL1*; *FASN*; *LPCAT1*; *MED27*). *LPCAT1* is of particular interest for two reasons. First, *LPCAT1* catalyzes the conversion of lysophosphatidylcholine (LPC) to phosphatidylcholine (PC, which comprises the majority of the total surfactant lipid composition [59]). The LPC to PC conversion plays a critical role in the maintenance of surfactant homeostasis via the lipid recycling cycle (Land’s cycle) [60]. Second, *LPCAT1* is connected to AT2 progenitor function and AT2-specific mitochondrial function deficiency [61, 62]. Knock-out of *LPCAT1* resulted in reduced oxygen conversion rate (OCR) in healthy AT2 cells, while overexpression of *LPCAT1* “rescues” oxygen conversion phenotypes in IPF AT2 cells [62]. Mitochondria play a key role in fueling surfactant lipid biosynthesis [63], connecting mitochondrial function to our observations of accumulated evidence for changes in surfactant lipid regulation in COPD/IPF. Together with our observations about *ATP11A*, these results support multi-omic evidence of switch-like alterations in lipid biosynthesis, which is critical for maintaining surfactant homeostasis in the lung, as a direction for further insight into the divergences of the lung toward COPD or IPF.

In a study of COPD patients, decreased surfactant levels were associated with worse lung function, and both PS and PE were markedly lower in COPD cases than in controls [64]; global dysregulation of surfactant lipids and proteins has been associated with COPD status, CT imaging metrics, and measures of lung function [65]. Changes in surfactant composition and surface tension properties have long been understood to be implicated in IPF, with lower surfactant levels observed in bronchoalveolar lavage (BAL) fluid in patients with IPF vs controls [66]. However, in cell culture models of fibrosis, type II alveolar epithelial cells exhibited an increased release of lipids, including PS, in response to injury along with reduced lipid reuptake and consequent accumulation of surfactant lipids in BAL fluid [67]. Importantly, these changes were apparent before the observation of fibrosis and thus may be causal [67]. In keeping with this prior literature, the switchlike behavior we observe in lipid metabolism-related genes may relate to decreases in surfactant in COPD and to changes in surfactant lipid homeostasis in IPF; further investigation is warranted.

The work presented here has several key strengths. The well-characterized LTRC cohort includes detailed physician-adjudicated phenotyping and high-quality assessment of important covariates such as ancestry (TOPMed ancestry PCs) and smoking history (methylation biomarkers). Methylation data are from the well-established Illumina EPIC array; we conducted our analyses using state-of-the-art software specific to this assay. This is the first paper to describe multi-omic switches for COPD and IPF, including the use of network methods to identify key epigenetic switches implicated in the divergence between COPD and IPF. The application of community detection to these networks provides a novel lens for discovering modules of CpGs whose collective switch-like behavior characterizes differences between COPD and IPF and identifying local hubs of these modules as potential targets for therapeutic intervention. Integrating transcriptomic data enables us to attain multi-omic mechanistic insights.

This study also has important limitations. Array-based methylation assays capture methylation only at certain genomic loci, biased towards genomic regions of interest[68]. We analyzed bulk data and relied on *in silico* deconvolution, which may not reflect cellular subtype differences (e.g., ciliated, alveolar type 1 or type 2 epithelial cells). Both COPD and IPF involve substantial remodeling of lung tissue; in both diseases, we observe disease-associated differences in cell proportion (Supplementary Figure E2b). Future single-cell methylation technology will permit cell type-specific insights. Controls in this study mostly had lung resection for lung nodules (benign, cancer) and therefore are not “healthy” controls. However, there was a concerted effort to obtain the samples as far away from the lesion as possible. The study consists of primarily white individuals, limiting generalizability to diverse populations. Future work exploring the relationship between ancestry, race, and ethnicity in the context of the COPD and IPF epigenetics is essential. This study lacks external validation with similar large-scale lung tissue validation cohorts. Future studies of lung tissue multi-omics in COPD and IPF are necessary to solidify the evidence for the molecular switches identified here. Finally, these data are cross-sectional; we cannot infer causality.

In sum, this work identified divergent epigenetic signatures of COPD and IPF. These findings implicate epigenetic “switch” regulation in the bifurcating etiology of COPD and IPF and highlight switch-like methylation of lipid-related genes as a possible mechanism for differences discriminating the two conditions. Our identification of epigenetic regulation of such genes in both diseases underscores the epigenetic plasticity of these mechanisms and highlights lipid-related genes (in particular, *LPCAT1* and *ATP11A*) for future mechanistic investigations and therapeutic innovation.

Postmatter

## Acknowledgements and funding

NHLBI TOPMed:

Lung Tissue Research Consortium Molecular data from the Trans-Omics in Precision Medicine (TOPMed) program was supported by the National Heart, Lung, and Blood Institute (NHLBI). DNA methylation for “NHLBI TOPMed: Lung Tissue Research Consortium” (phs001662) was performed at the Northwest Genomics Center (HHSN268201600032I). Core support including centralized genomic read mapping and genotype calling, along with variant quality metrics and filtering were provided by the TOPMed Informatics Research Center (3R01HL-117626-02S1; contract HHSN268201800002I). Core support including phenotype harmonization, data management, sample-identity QC, and general program coordination were provided by the TOPMed Data Coordinating Center (R01HL-120393; U01HL-120393; contract HHSN268201800001I). We gratefully acknowledge the participants who provided biological samples and data for TOPMed.

This work was supported by the National Institutes of Health under the following awards:

- KHS was supported by P01HL114501 and T32HL007427 and K25HL175222.
- JHY was supported by K08HL146972.
- BDH was supported by R01HL162813, R01HL155749, R01HL160008, and U01HL089856. BDH was also supported by a Research Grant from the Alpha-1 Foundation.
- EKS was supported by P01HL114501.
- JQ was supported by R35 CA220523, P01HL114501, and R01HG011393.
- DLD was supported by K24HL171900, R01HG011393, R01HL178032 and P01HL114501.

The content is solely the responsibility of the authors and does not necessarily represent the official views of the National Institutes of Health.

## Data availability statement

LTRC genotyping for the generation of ancestry PCs and LTRC DNA methylation data are available on dbGaP with accession number phs001662.v4.p2. dbGaP can be accessed at https://www.ncbi.nlm.nih.gov.

## Human subjects statement

All subjects provided written informed consent per LTRC protocol. The study was approved by the Institutional Review Board of each participating institution.

## Supplementary Material

### 1. Supplementary Material

#### 1.1 Study Population

The Lung Tissue Research Consortium (LTRC) is a centralized biobank established by the National Heart, Lung, and Blood Institute of the National Institutes of Health in the United States (https://biolincc.nhlbi.nih.gov/studies/ltrc/) [1, 2]. From 2005 to 2019, the LTRC enrolled 4,486 subjects and obtained lung tissue samples from 3,333 of these individuals during medically indicated lung resection procedures. The study was approved by institutional review boards at each participating institution, and written informed consent was provided by all subjects per LTRC protocol. Here, we analyze methylation data generated through the Trans-Omics for Precision Medicine program from 1,063 LTRC participants.

#### 1.2 Assessment of lung disease phenotypes

Control, COPD, and IPF phenotypes were assessed using spirometry, clinical diagnoses, and lung tissue pathology. Controls were defined by normal spirometry (forced expiratory volume in one second (FEV_1_) ≥ 80% predicted and FEV_1_/forced vital capacity (FVC) ≥ 0.7) without a clinical diagnosis (by LTRC site principal investigator) of sarcoidosis or idiopathic pulmonary fibrosis (IPF) and without a pathologic diagnosis of interstitial lung disease (ILD)/ idiopathic interstitial pneumonia (IIP), sarcoidosis, constrictive bronchiolitis, cellular hypersensitivity pneumonitis, eosinophilic granulomas, or diffuse alveolar damage (DAD). COPD cases were defined by abnormal spirometry (FEV_1_ < 80% predicted and FEV_1_/FVC < 0.7) without a clinical diagnosis (by LTRC site PI) of sarcoidosis or IPF and without a pathologic diagnosis of ILD/IIP, sarcoidosis, constrictive bronchiolitis, cellular hypersensitivity pneumonitis, eosinophilic granulomas, or DAD. IPF was defined by an expert summative clinical diagnosis, in most cases corresponding to pathology.

#### 1.3 Covariate assessment

EWAS models were adjusted for age, sex, cigarette smoke exposure, ancestry, estimated cell type composition, and plate, as we observed batch effects associated with plate (Supplementary Figure E6). Age was assessed based on the date that lung tissue was obtained. Biological sex was verified by cross-referencing with whole-genome sequencing data and allosomal methylation and matched with the self-report. Exposure to cigarette smoke was assessed according to methylation of the probe cg05575921 in the aryl hydrocarbon receptor repressor gene *AHRR*. Methylation of *AHRR* at the cg05575921 locus (“AHRR methylation”) is a reliable biomarker for smoking status; current smoking and higher smoking intensity are associated with lower AHRR methylation in blood[3]. We verified the relationship between smoking and AHRR methylation in our data by cross-referencing with self-reported smoking status where available (Supplementary Figure E2).

Twenty ancestry principal components (ancestry PCs) were estimated from the complete set of TOPMed genetic data using Locating Ancestry from SEquence Reads (LASER) [4–6]. We included the first ten of these ancestry PCs in the EWAS models in an effort to parsimoniously capture substantial variation due to ancestry.

#### 1.4 Preprocessing of methylation array data

Preprocessing of methylation array data followed a standard workflow using well-established methylation array pipelines annotated against the Illumina B4 manifest (Supplementary Figure E5). Analyzed probes were restricted to only those present on the B4 manifest. Probes with a detection p-value > 0.05 in more than 25% of samples, non-CpG probes, probes with known SNPs at the CpG site or at the single-nucleotide extension with minor allele frequency >= 0.05[7–9], cross-reactive probes[10, 11], and probes with less than 3 bead numbers in at least 20% of the samples were removed from the analysis. The minfi R package was used for noob-adjustment[7, 8, 12], the rcp algorithm was used to correct for probe type bias[13], and functional normalization was performed using the EnMix R package[14].

Inspection of the principal components of a random sample of N = 5000 probes identified a batch effect associated with the plate on which each sample was run (Supplementary Figure E6a-b). Because plate and phenotype (control, COPD, IPF) showed some association in our dataset, we adjusted for plate in the EWAS models rather than performing batch correction in order to avoid inducing artificial effects in the batch correction process [15]. Methylation beta values were converted to M values using the base-2 logit function, yielding continuous values on the real line; use of M values in regression models is preferred due to heteroscedasticity in beta values observed among sets of CpGs [16].

#### 1.5 Estimation of cell type composition

Cell type composition was estimated using EpiSCORE [17]. In lung tissue, EpiSCORE estimates seven cell types: endothelial cells, epithelial cells, granulocytes, lymphocytes, macrophages, monocytes, and stromal cells. Because these estimates are compositional (sum to one), it is necessary to choose a baseline cell type to exclude from the model. We set endothelial cells as the baseline and adjusted for the other six cell types in the EWAS model.

#### 1.6 Statistical methods for EWAS

To identify associations between methylation and lung disease phenotype, we performed two separate epigenome-wide association studies (EWAS), one comparing COPD to controls and one comparing IPF to controls. EWAS models consisted of linear regression of methylation M-values against an indicator of control vs. lung disease (COPD or IPF). Models were adjusted for age, sex, *AHRR* methylation at cg05575921, ancestry principal components, estimated cell type composition, and plate. Throughout the manuscript, p-values less than machine zero in R are reported as < 2.2e-16.

#### 1.7 Lung tissue replicates

The LTRC lung tissue methylation data consist of 1708 samples, including 131 spatial replicates from 90 subjects in which multiple lung tissue samples were obtained from different regions of the lung in the same patient and 21 technical replicates from 20 subjects in which multiple assays were run on the same sample (Figure 1). Exploratory analyses indicated high correlation between both spatial and technical replicates (Supplementary Figure E7). In lung, the average correlation between spatial replicates was 0.989 and the average correlation between technical replicates was 0.992. Based on these analyses, we selected one sample at random for any participant with either type of replicate. We excluded participants missing ancestry PCs (N=78), failing methylation QC (N=37), or not classified as control, COPD, or IPF (N=412); the remaining dataset consisted of 1029 samples.

#### 1.8 Test statistic post-processing

EWAS test statistics were corrected for bias and inflation using the R package BACON[18]. We implemented two options to account for multiple hypothesis testing when assessing significance. First, as a conservative measure, we established a genome-wide significance threshold of 6.3*10^-8 based on a Bonferroni correction of N=790,856 tests (the number of probes modeled in both lung and blood) at significance level 0.05. As a second, less conservative, measure, BACON p-values were adjusted using the Benjamini-Hochberg FDR[19] with the R function p.adjust (method=“bh”) and any probes associated with COPD or IPF at FDR < 0.05 were considered significant.

Of particular interest are the set of probes significant in both COPD and IPF, which we refer to as *overlap probes*, and the subset of overlap probes with opposite direction of effect, which we refer to as *switch probes*.

#### 1.9 Regional analysis with DMRcate

DNA methylation data typically display high spatial correlation, and it is plausible that methylation of a genomic region is functionally more relevant than methylation of individual CpGs[20]. Moreover, when comparing across conditions (control, COPD, PF) or tissues (lung, blood), there may be probe-by-probe differences in a genomic region in which regional methylation is concordant, or vice-versa. To investigate these possibilities in our data, we performed regional differential methylation analyses using DMRcate[21]. As in the EWAS, two differential analyses were conducted: (i) a comparison of COPD vs. control, (ii) a comparison of IPF vs. control.

We applied the dmrcate function to the BACON-corrected t-statistics from each EWAS to detect differentially methylated regions (DMRs), using the recommended bandwidth parameter lambda=1000 and the scaling parameter C=2 for the kernel smoothing algorithm and requiring a minimum of 2 CpGs to define a DMR. The recommended bandwidth parameter is based on empirical performance on the 450k array[21].

Comparison of DMRs is more complex than comparison of DMPs in that one must select a definition of regions which “match” between conditions. We explored three types of matches. First, we considered two DMRs to be overlapping if they were within a 20-basepair window up or downstream of each other. We used the R package GenomicRanges[22] to identify these overlaps (parameters: minoverlap=0 and maxgap=20). Second, we restricted our definition of shared DMRs to exact matches, i.e., DMRs that share the same genomic coordinates. Third, we explored matches at the gene level. For each DMR, DMRcate identifies a list of genes overlapping the region. We looked for concordance between these gene sets across conditions.

#### 1.10 ORA of differentially methylated probes (DMPs) and switch DMPs

To distill biological meaning from the EWAS hits, we performed over-representation analyses (ORA) using the gometh function of the R package missMethyl [23]. This function uses the HG19 genome assembly. We conducted two separate ORAs considering (1) hits in COPD, and (2) hits significant in IPF. We also conducted a third ORA considering the set of 1,091 lung switch probes. Based on statistical properties of the gometh function, we report nominal p-values as in the original missMethyl paper[23]. Pathways with p < 0.05 were deemed enriched. We investigated the KEGG legacy pathways gene set collection [24–26], which is directly integrated into missMethyl.

#### 1.11 ORA of genes overlapping switch differentially methylated regions (DMRs)

We performed an over-representation analysis on the set of unique genes annotated to the switch DMRs in lung using the fora function of the fgsea R package(Korotkevich et al., 2016). For this analysis, we used a set of N=186 KEGG legacy gene sets[24–26] downloaded from the Molecular Signatures Database[27, 28] (c2.cp.kegg_legacy.v2023.2.Hs.symbols.gmt). The background universe of genes was defined as the set of N=47,621 unique genes on the EPIC array as annotated by the EPIC manifest mapped to the HG38 annotated with GENCODEv36, available at https://zwdzwd.github.io/InfiniumAnnotation. The set of genes overlapping the switch DMRs in COPD may vary from those overlapping the switch DMRs in IPF based on the definition of regional overlap described in Section 4.7; therefore, we considered the union of these sets to be representative of the gene-level switch DMR signal. We note that this approach does not account for potential bias due to the variation in the number of probes per gene; however, such bias may be mitigated to some extent by the regional analysis, which combines the signals of genomically proximal probes.

#### 1.12 Graphical modeling of switch probes in lung

Gaussian graphical models (GGMs, also known as partial correlation networks) were used to interrogate network-level relationships among switch probes. To estimate GGMs, we first regressed the methylation data against the same covariates used in the EWAS model (age, sex, AHRR methylation, ancestry, estimated cell type composition, and plate). Using the residuals from these regressions, we next applied the graphical lasso[29] as implemented in the R package huge[30]. The extended Bayesian Information Criterion (eBIC)[31] was used to select the penalty parameter, with the tuning hyperparameter gamma set to zero, encouraging a denser network. The influence of individual nodes in this network was quantified using hub centrality[32], calculated using the hits_scores() function in the R package igraph[33]. A GGM consists of positive and negative edge weights representing partial correlations; since both types of associations are equally important in our investigation, we took the absolute value of edge weights prior to calculating hub score.

To perform community detection, we applied Louvain clustering using the cluster_louvain algorithm as implemented in igraph[33, 34]. The resolution parameter was set to the default value (resolution = 1). Because Louvain clustering has a stochastic component, we used a random seed to ensure reproducibility. In a sensitivity analysis, we assessed stability of the detected communities by rerunning the community detection 100 times and calculating the normalized mutual information between pairs of community structures using the igraph function compare() (Supplementary Figure E8).

To identify local hubs within each community, we applied the hits_scores() function to the subgraph of each community. To explore the stability of local hub selection, we repeated this process in the 100 simulated community structures described above and counted how many times each CpG was selected to be a local hub (Supplementary Table E8d).

To perform enrichment analysis, we used missMethyl [23] as described in the section “ORA of differentially methylated probes (DMPs) and switch DMPs”.

#### 1.13 Methods for assessing differential methylation at the gene level (differentially methylated genes; DMGs)

Because the number of probes per gene on the EPIC array is highly variable, we conducted a statistical test to identify differentially methylated genes (DMGs) based on the significance of the number of DMPs per gene. We assumed a null model in which the total number of DMPs observed in the EWAS are evenly distributed across the set of probes on the array. With this assumption, the number of DMPs observed at each gene *k* is a hypergeometric distribution: we observe a set of *m(k)* EWAS hits in a random sample of size *n(k)* from the set of N total probes where *n(k*) is the total number of probes on the EPIC array mapping to gene *k* and sampling is without replacement. Based on this distribution, we used a Fisher’s exact test to calculate the one-tailed probability of observing more than *m(k)* EWAS hits on gene *k* [35]. We corrected for multiple testing by calculating FDR using the p.adjust() function in R (method = “BH”). A gene was considered to be a DMG if the Fisher’s exact test yielded an FDR < 0.05.

#### 1.14 Details of genes previously associated with COPD and IPF

To assess DNA methylation signals among genes that have previously been reported to be implicated in COPD and IPF, we began by compiling a list of genes from relevant literature. We manually curated 15 gene sets consisting of 132 unique genes in total (Supplementary Table E9a). For each gene, probe annotations were obtained from the EPIC manifest mapped to the HG38 annotated with GENCODEv36, available at https://zwdzwd.github.io/InfiniumAnnotation.

In total, 6,399 probes were annotated to the 132 genes from the literature. Gene sets included a set of genes related to telomere function (“telomere_genes”; [36]), the shelterin complex (“shelterin_genes”; [37]), alpha-1 antitrypsin deficiency (“aat_genes”), MUC5B (“muc5b_genes”), surfactants (“surfactant_genes”), a selection of genes involved in the Hippo signaling pathway (“hippo_genes”;[38–40], switch genetic variants from previous GWAS of COPD and IPF (“hobbs_switch_genes”; [41], “sakornsakolpat_switch_genes”; [42], “hobbs_2017_gwas_snp_genes”; [41]), genes previously implicated in the divergent mechanisms of COPD and IPF (“chilosi_switch_genes”; [43]), genes encoding proteins used in an integrative protein-protein interaction (PPI) network analysis of COPD and IPF (“halu_copd_seed_genes”, “halu_ipf_genes”, “halu_2019_module_overlap_genes”;[44]), and sets of differentially expressed COPD and IPF genes with previous GWAS associations (“ghosh_copd_gwas_de_genes”, “ghosh_ipf_gwas_de_genes”;[1]),

#### 1.15 Multi-omic integration with gene expression data

Previous differential expression analyses in LTRC lung tissue identified 5010 genes differentially expressed between COPD and control and 11,454 genes differentially expressed between IPF and control (FDR < 0.01) ([1] “Ghosh results”). To perform a targeted integration of our EWAS results with the transcriptomic results, we began with the set of genes annotated to our switch probes based on the EPIC manifest mapped to the HG38 annotated with GENCODEv36, available at https://zwdzwd.github.io/InfiniumAnnotation. The Ghosh results are annotated with gene names based on GENCODEv29. To ensure consistency across GENCODE versions, we first searched for our GENCODEv36 gene names in the Ghosh results. For any of our genes that were not matched, we then looked for alternative names in GENCODEv29 based on matching on Ensemble transcript IDs (ENST).

We then took the set of switch probes annotated to genes matching differentially expressed genes in the Ghosh results and examined the relationship between our EWAS results and the log fold change (logFC) of the gene expression in COPD vs. control and in IPF vs control. Using this mapping, we identified a set of “canonical epigenetic switch probes” (CES probes) as defined by the following three criteria (see also flowchart in Supplementary Figure E9):

1. Probe is a switch probe, exhibiting significant differential methylation in both COPD vs. control and in IPF vs. control (FDR < 0.05) with opposite directions of effect in COPD and IPF.
2. Probe is annotated to a gene described in the Ghosh results as differentially expressed in both COPD vs control and IPF vs control (FDR < 0.01), with opposite directions of effect in COPD and IPF.
3. Probe-gene relationship exhibits what we call “canonical” epigenetic behavior: higher methylation is associated with lower expression and vice-versa.

To interrogate the function of genes annotated to the CES probes, we used Enrichr [45–47] to map these genes to the BioPlanet pathways [48].

### 2. Supplementary Results and Discussion

#### 2.1 Probes of interest in graphical models

2.1.1 cg03977586 methylation is negatively associated with COPD (effect: −0.21, FDR = 0.003) and positively associated with IPF (effect: 0.43, FDR = 0.02). This CpG is located on chromosome 12 in an exon of *WASHC4*, downstream of a predicted ZNF680 binding site and upstream of binding sites for a number of JUNx:FOSx (activating protein 1, or AP-1) dimers.

2.1.2. cg07262457 methylation is positively associated with COPD (effect: 0.08, FDR=0.025) and negatively associated with IPF (effect: −0.18, FDR=0.020) This CpG is located on chromosome 3 in a region classified by ENCODE as a CTCF-bound distal enhancer-like signature (dELS).

2.1.3 cg17200083 stands out for its high significance in the IPF EWAS (effect: - 0.56, FDR = 8.63e-12). This probe is located on chromosome 1 in an intron of *PLXNA2*, near the 3’ end of a predicted binding site for *ZNF282*.

2.1.4 cg09180057 has a strong positive association with COPD (effect: 0.36, FDR < 2.2e-16) and a high hub score (0.78). Methylation of cg09180057 is positively associated with IPF (effect:-0.51, FDR = 7.49e-7), with a moderate hub score (0.63). This probe is located in a *FOXN3* intron (chr14), falling downstream of a predicted binding site for *SOX4* and upstream of a predicted binding site for *FOXD2* as well as downstream of an ENCODE dELS (EH38E1734456).

#### 2.2 Wnt, Notch, and EMT-*related evidence*

Dysfunction of the epithelial-mesenchymal transition (EMT) is an established component of both COPD and IPF[49]. Wnt signaling and Notch signaling are two key EMT-related developmental pathways for which switch-like behavior has been observed between COPD and IPF, with decreased Wnt/Notch signaling observed in COPD and increased Wnt/Notch signaling in IPF[43, 50]. We observed epigenetic evidence related to both of these pathways in our investigations of lung tissue, as well as in other EMT-related genes and processes including JAK-STAT and Hippo signaling.

The genome wide-significant switch DMP cg09180057 mapped to *FOXN3*, which has been shown in cancer to suppress Wnt/beta-catenin signaling[51, 52]. In IPF, the KEGG gene set hsa04310:Wnt signaling pathway was overrepresented among DMPs, while in COPD, it was not, supporting this switch hypothesis. In the switch DMRs, the KEGG gene set hsa04330:Notch signaling was overrepresented, driven by the genes *CTBP1*, *DLL1*, *LFNG, MAML2*, *NOTCH1,* and *NOTCH4*. *NOTCH1* was one of the genes identified in the literature as a possible switch mechanism between COPD and IPF[43], and we observed *NOTCH1* to be differentially methylated at the gene level in IPF but not in COPD (Table 4). The KEGG Notch signaling pathway itself was overrepresented in DMPs in IPF but not COPD, further suggesting a switch mechanism.

In addition to Wnt/Notch, we noted differential methylation in other EMT-related genes and processes. The genome wide-significant switch DMP cg02845735 mapped to *FOXP1*, which has demonstrated an inverse association with the EMT via E-cadherin and snail[53]. DMP enrichment analyses implicated JAK-STAT in COPD but not IPF and Hippo signaling in IPF but not COPD (Supplementary Table E5). The gene *LMNA* overlapped a top switch DMR (Table 3); *LMNA* has been implicated in reversal of the EMT in a human cell culture model of non-small cell lung cancer (NSCLC) (HCC827 cells)[54]. In the switch probe graphical models, probes mapping to *NTN1* were local hubs in both the COPD and IPF networks (Supplementary Table E8b). *NTN1* has been shown to inhibit TGFbeta1-induced EMT in a human cell culture model of NSCLC (A549 cells) [55]. Two of the genes in our literature scan arose in relation to the EMT: *ATP11A* and *AKAP13*. *ATP11A*, differentially methylated in both COPD and IPF (Table 4), has been shown to regulate Hippo signaling in gastric cancer cells, resulting in increased EMT[56]. *AKAP13*, for which we observed gene level differential methylation in COPD but not IPF (Table 4), has been shown to regulate the Hippo signaling pathway in granulosa cells [57]; although the setting of ovarian pathobiology differs from that of the lung, it is possible that *AKAP13* serves a similar regulatory role in COPD, where the Hippo signaling pathway has been implicated[58]. A role for *AKAP13* in the EMT via TGF-beta activation has also been proposed[59].

#### 2.3 Literature comparison

##### 2.3.1 Details of genes previously associated with COPD and IPF in the literature that are differentially methylated in our study

*AKAP13* was significantly differentially methylated in COPD (FDR = 0.028) and not in IPF (FDR = 0.255). *AKAP13* has been previously connected to IPF in the literature and was chosen for analysis here based on its differential mRNA expression, with *AKAP13* levels elevated in IPF cases vs. controls[1, 60]. A variant near *AKAP13* (rs62025270) was identified by Allen et al. (2017) as a novel GWAS association for IPF. However, this variant is relatively common compared to the rarity of IPF: about 1 in 3 individuals of European descent carry the risk allele, and it is posited that other mechanisms complement genetic variation in determining the course towards IPF[61]. Although not meeting FDR significance at the gene level, our identification of 17 DMPs mapped to *AKAP13* suggest that epigenetic regulation could be one such mechanism. For most *AKAP13* probes, methylation is lower in IPF cases than controls (Figure 5), consistent with the increased mRNA expression observed in previous studies [1, 60].

In contrast to its involvement in IPF, *AKAP13* has not been extensively connected to COPD in the literature, and its differential methylation in our dataset represents a novel link. A proposed role for *AKAP13* in the EMT is discussed in the main manuscript (Section 3.1.2).

NOTCH1 was also included in our literature review along with several other Notch genes, based on evidence for opposing roles of Notch signaling in COPD and IPF (Chilosi et al., 2012; Li et al., 2015). We observed NOTCH1 to be differentially methylated with generally higher methylation in IPF vs. controls; however, it was not differentially methylated in COPD. This difference in epigenetic regulation could be part of determining the opposing roles of Notch signaling in COPD and IPF. In IPF, increased Notch1 signaling has been characterized as pro-fibrotic and associated with type II alveolar epithelial cell proliferation as well as surfactant dysfunction[62]. We observed increased methylation in *NOTCH1* in IPF, which is typically associated with decreased expression; however, promoter vs. gene body methylation often have opposing effects. In COPD, ERK signaling downstream of *NOTCH1* has been shown to regulate promoter methylation of mitochondrial transcription factor A (mtTFA) in response to cigarette smoke exposure; it is possible the epigenetics of COPD are manifested via ERK signaling rather than in methylation of *NOTCH1* itself[63].

THSD4, included in our literature review as a GWAS hit for lung function in COPD [41, 64], was differentially methylated in both COPD and IPF with some switch-like behavior (Table 4, Figure 5). We found one switch probe, cg05673257 annotated to THSD4, located in the south shelf of a CpG island. The COPD effect for this probe was 0.11 (FDR = 0.039) and the IPF effect was −0.265 (FDR = 0.016). A second THSD4 probe, cg17882867, was also of interest as it was previously shown to be differentially methylated in COPD lung tissue, with higher methylation in those with COPD vs. controls (Morrow et al., 2016). We validated this result in our dataset, observing higher methylation at this probe site in COPD vs controls (effect = 0.15, FDR = 0.005). Although genetic variation in THSD4 has not yet been implicated in IPF risk, a SNP in a THSD4 intron (rs12899618) was associated with FEV1/FVC in a large study of over 20,000 individuals, including people with and without COPD [64]. Interestingly, FEV1/FVC typically decreases in individuals with COPD and increases in individuals with IPF[65], supporting a switch-like phenotype consistent with the switch-like epigenetic signature we observed in THSD4.

##### 2.3.2 Telomeres

Premature telomere shortening is a well-established component of the pathogenesis of COPD and IPF[43]. While mutations in telomere-related genes (such as *TERT* and *TERC*) have been implicated in both diseases [66], such mutations are absent in most patients[36]. Epigenetic mechanisms may explain the discrepancy between the relative infrequency of *TERT/TERC* mutations and the pervasiveness of short telomeres in COPD and IPF. We find support for this hypothesis: we observed differentially methylated probes in or near the telomere-related genes *NAF1, PARN*, *RAP1A, RAP1B, RTEL1, TERF1, TERF2, and TPP1*. Notably, these genes encode four of the six proteins comprising the shelterin complex (TRF1, TRF2, RAP1, TPP1), suggesting epigenetic changes in shelterin function as a possibly orthogonal mechanism to genetic variation in genes encoding telomerase-production machinery.

We found differential methylation in both COPD and IPF at several CpG sites mapping to *RTEL1*. In lung tissue, these probes generally had lower methylation levels in both COPD and IPF relative to healthy controls. This concordance of effect direction suggests that epigenetic control of *RTEL1* may be common to both diseases. Additionally, in COPD, one DMR mapped to *RTEL1* (chr20:63,682,383-63,682,861); mean methylation in this region was lower in COPD than in controls.

We also observed DMPs mapping to *PARN* with primarily lower methylation in IPF cases as compared to controls and null effects or higher methylation in comparing COPD and controls. These discordant directions of effect suggest *PARN* may be involved in processes that lead injured lung tissue along the path to either COPD or IPF. One *PARN* probe of particular interest because of its switch-like behavior is cg00463957 (chr16). This probe lies in an enhancer classified as a CTCF-bound dELS.

The remaining telomere-related genes, *RAP1A, RAP1B, TERF1, TERF2,* and *TPP1,* are related to the shelterin complex and suggest epigenetic changes in shelterin function as a possibly orthogonal mechanism to genetic variation in genes encoding telomerase-production machinery. We found two DMPs mapped to *TPP1* in lung tissue of individuals with IPF (Supplementary Table E9a) as well as a DMR overlapping *TPP1* (Supplementary Table E3). Additionally, the KEGG *RAP1* signaling pathway was nominally overrepresented among DMPs in both COPD and IPF lung tissue (Figure 3).

## Supplementary Figures

**Supplementary Figure 1:**
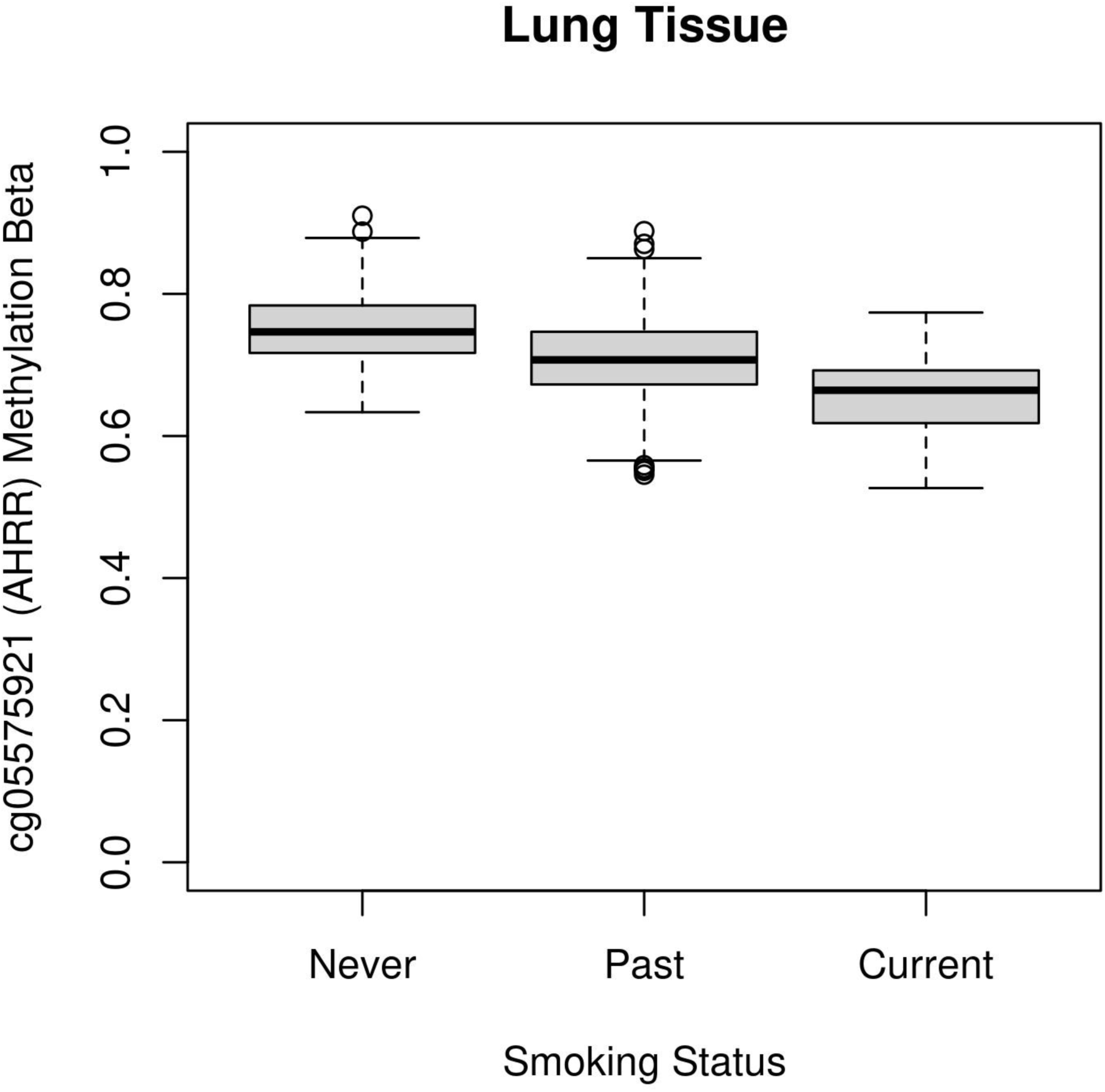
AHRR methylation by self-reported smoking status in lung

**Supplementary Figure 2a:**
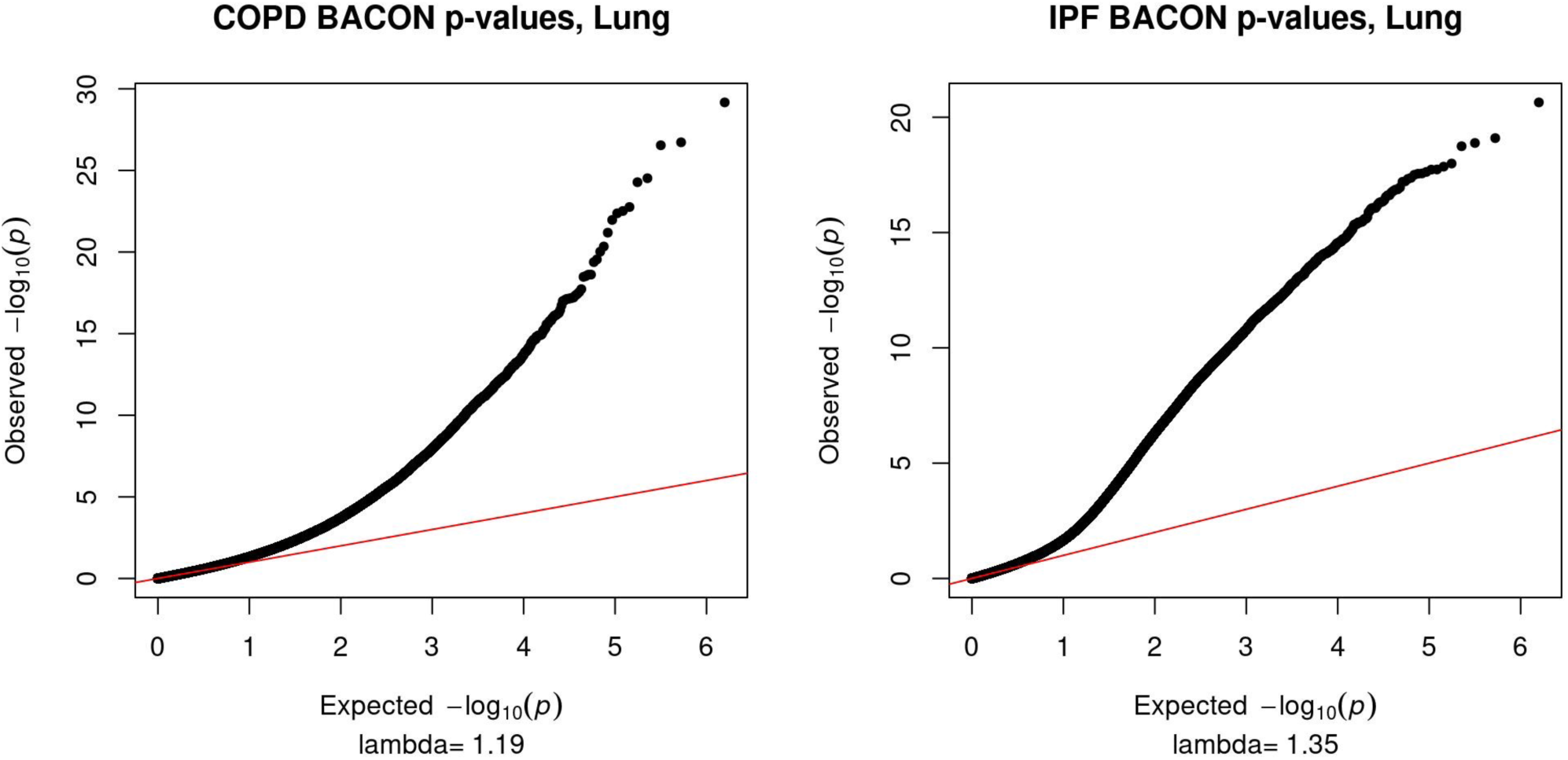
QQplots and genomic inflation factors of BACON-adjusted p-values in COPD and IPF.

**Supplementary Figure 2b:**
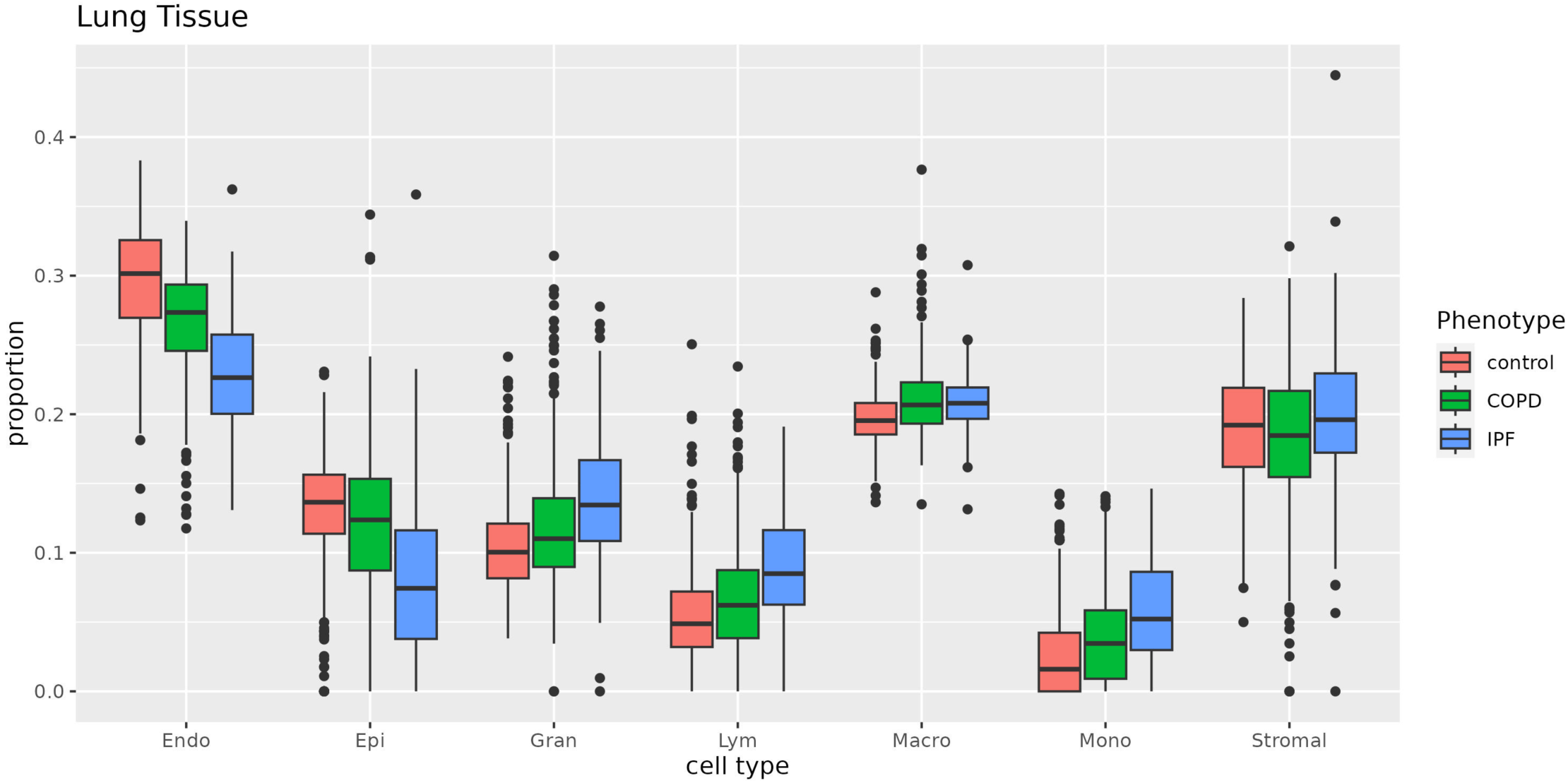
Disease-associated differences in estimated cell type composition.

**Supplementary Figure 3:**
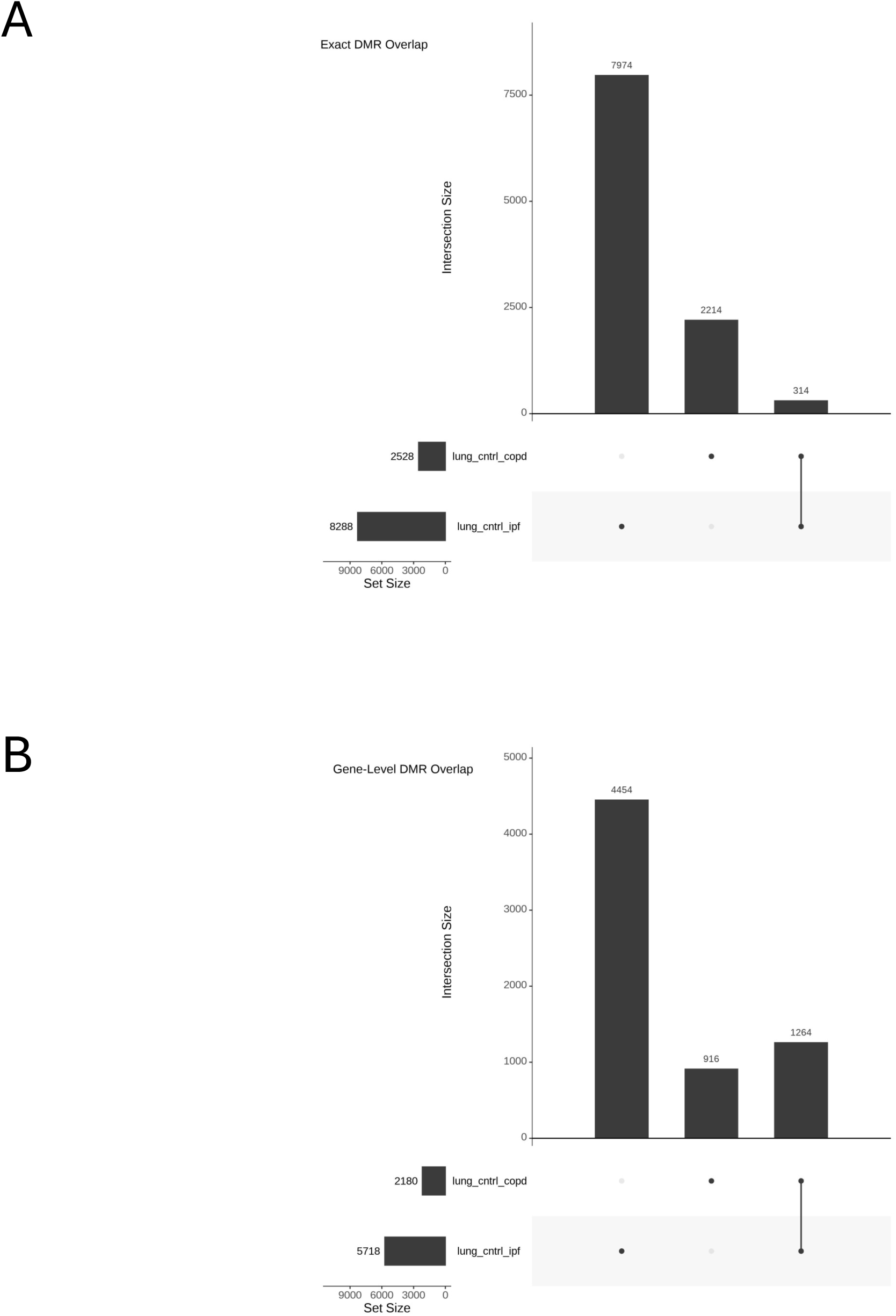
UpSet plots for DMR overlap between the COPD and IPF lung tissue EWAS. (a) Exact overlap, defined as identical genomic coordinates of the DMR. (b) Gene-level overlap, defined as overlaps of genes annotated to each DMR by DMRcate.

**Supplementary Figure 4:**
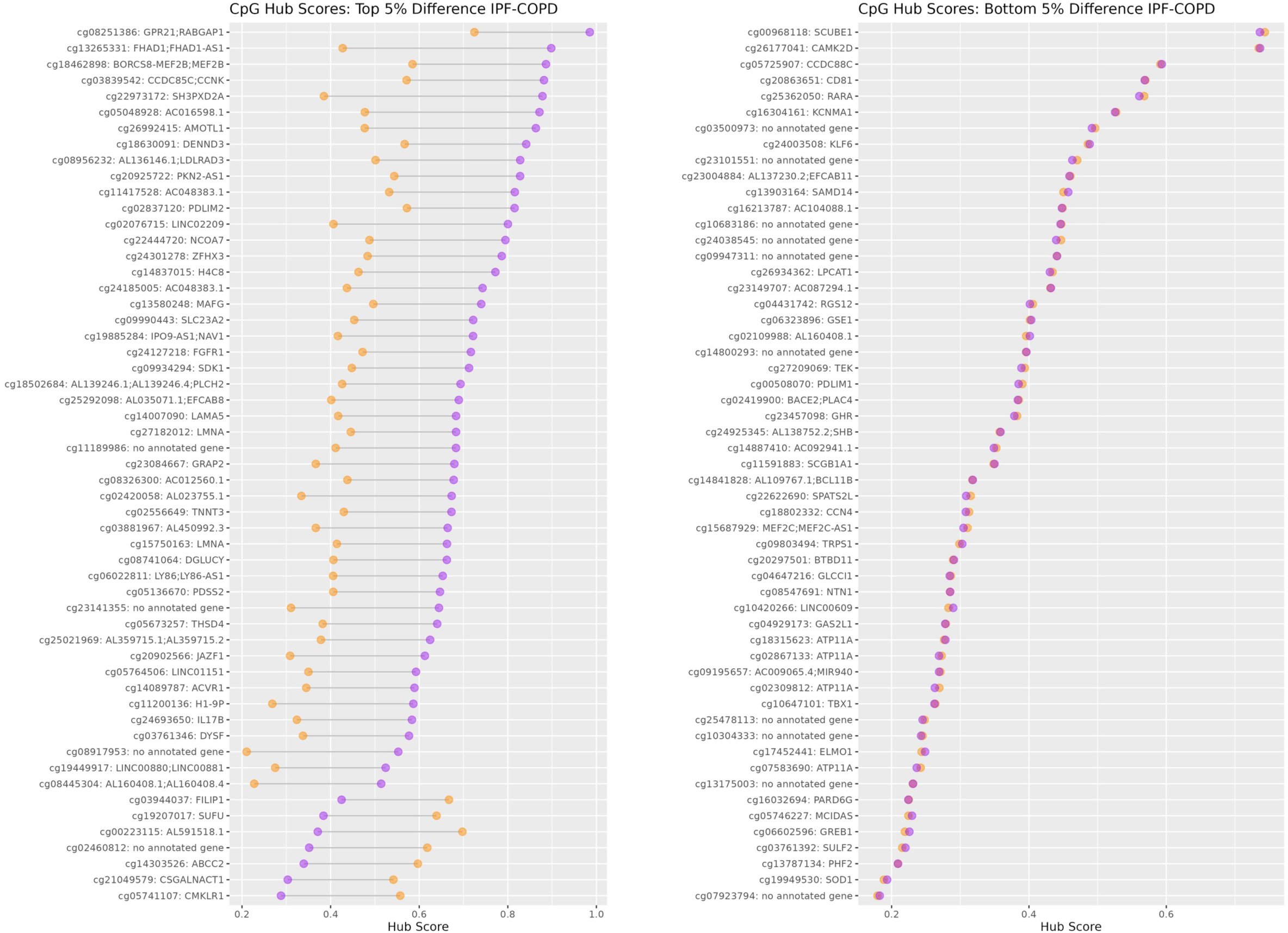
Comparison of switch probe hub score in COPD and IPF. (a) Switch probes with the largest difference in hub score between COPD and IPF. (b) Switch probes with the smallest difference in hub score between COPD and IPF.

**Supplementary Figure 5:**
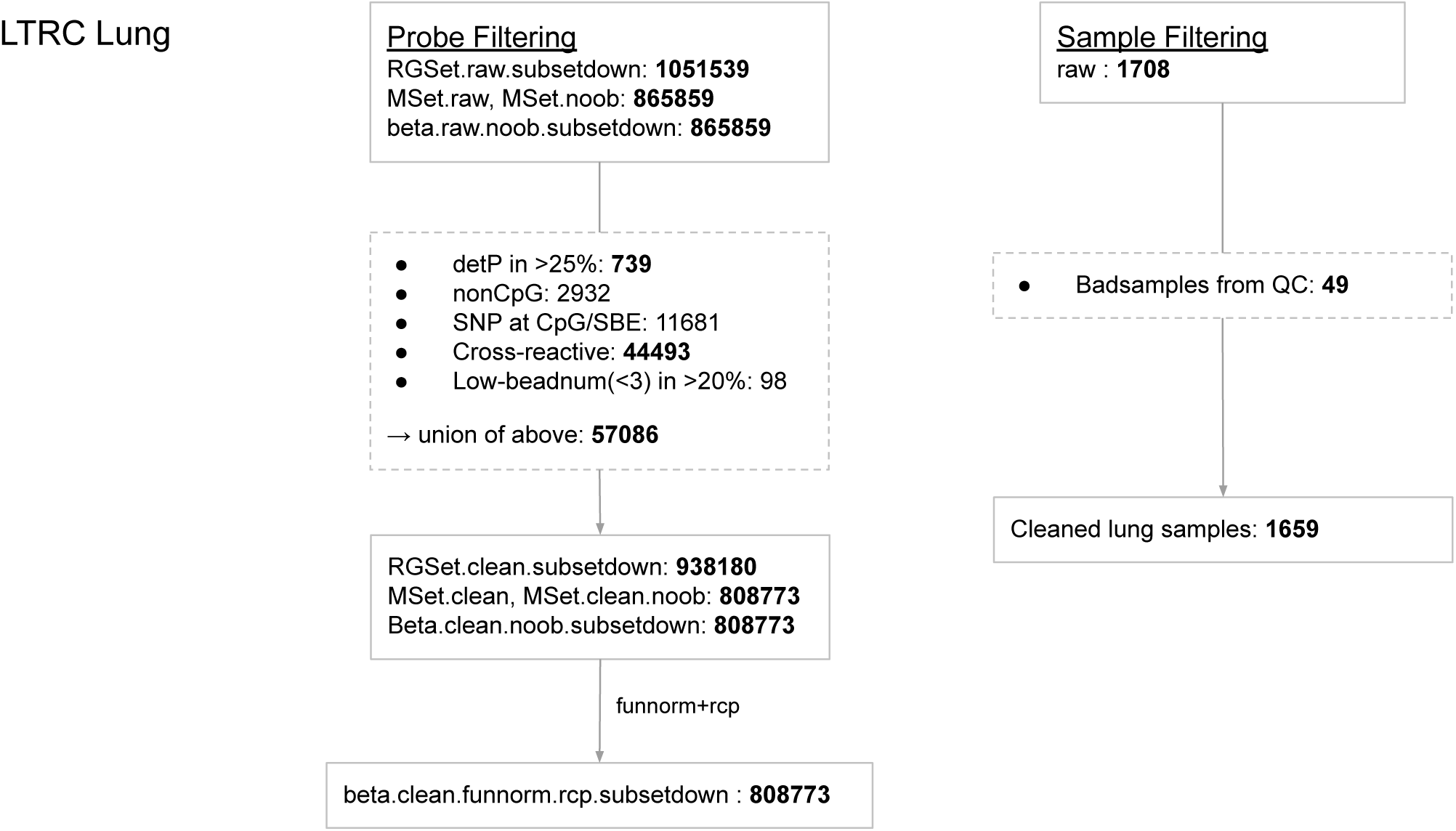
Workflow for filtering probes and samples during the data cleaning process.

**Supplementary Figure 6:**
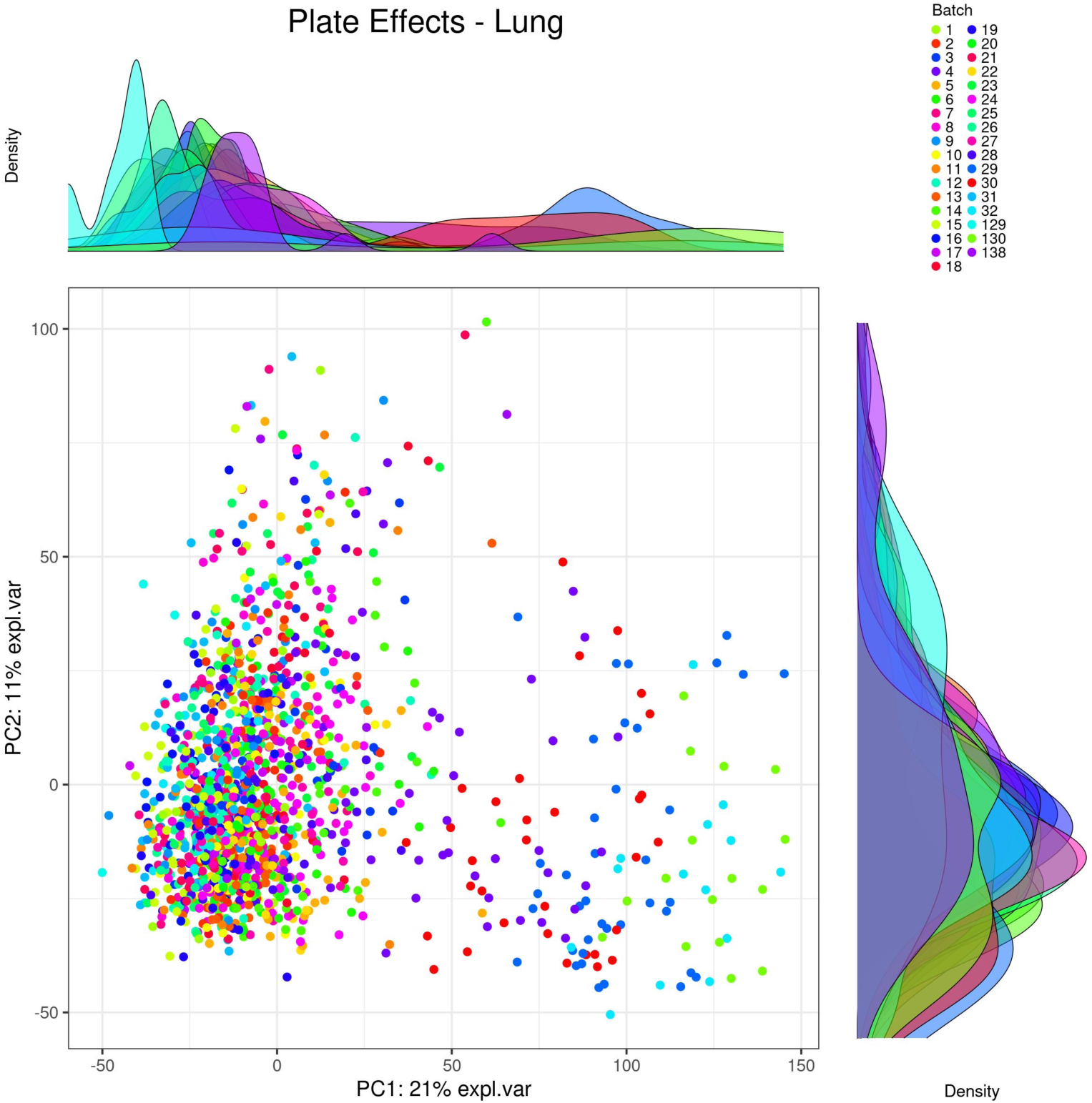
Diagnosis of batch effect in the Plate variable. A random subsample of the array probes were used to calculate principal components.

**Supplementary Figure 7:**
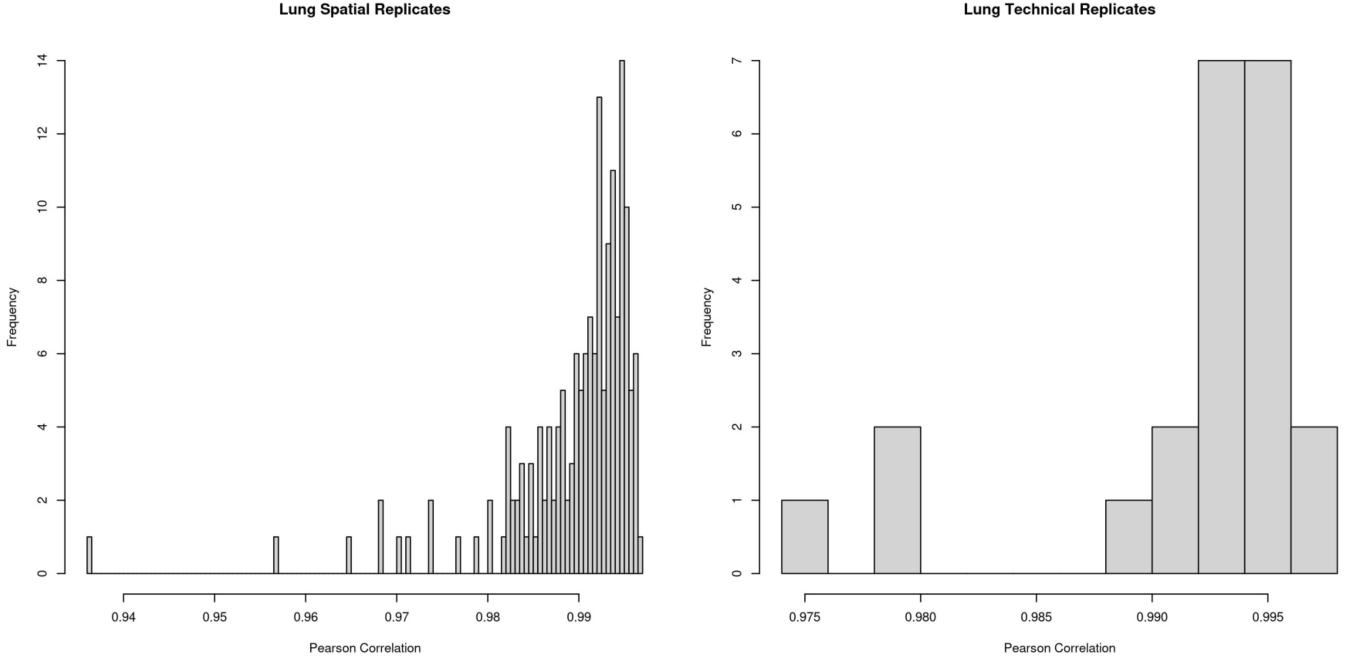
Histogram of pairwise Pearson correlation between spatial (multiple lung tissue samples from different lobar locations) and technical replicates.

**Supplementary Figure 8:**
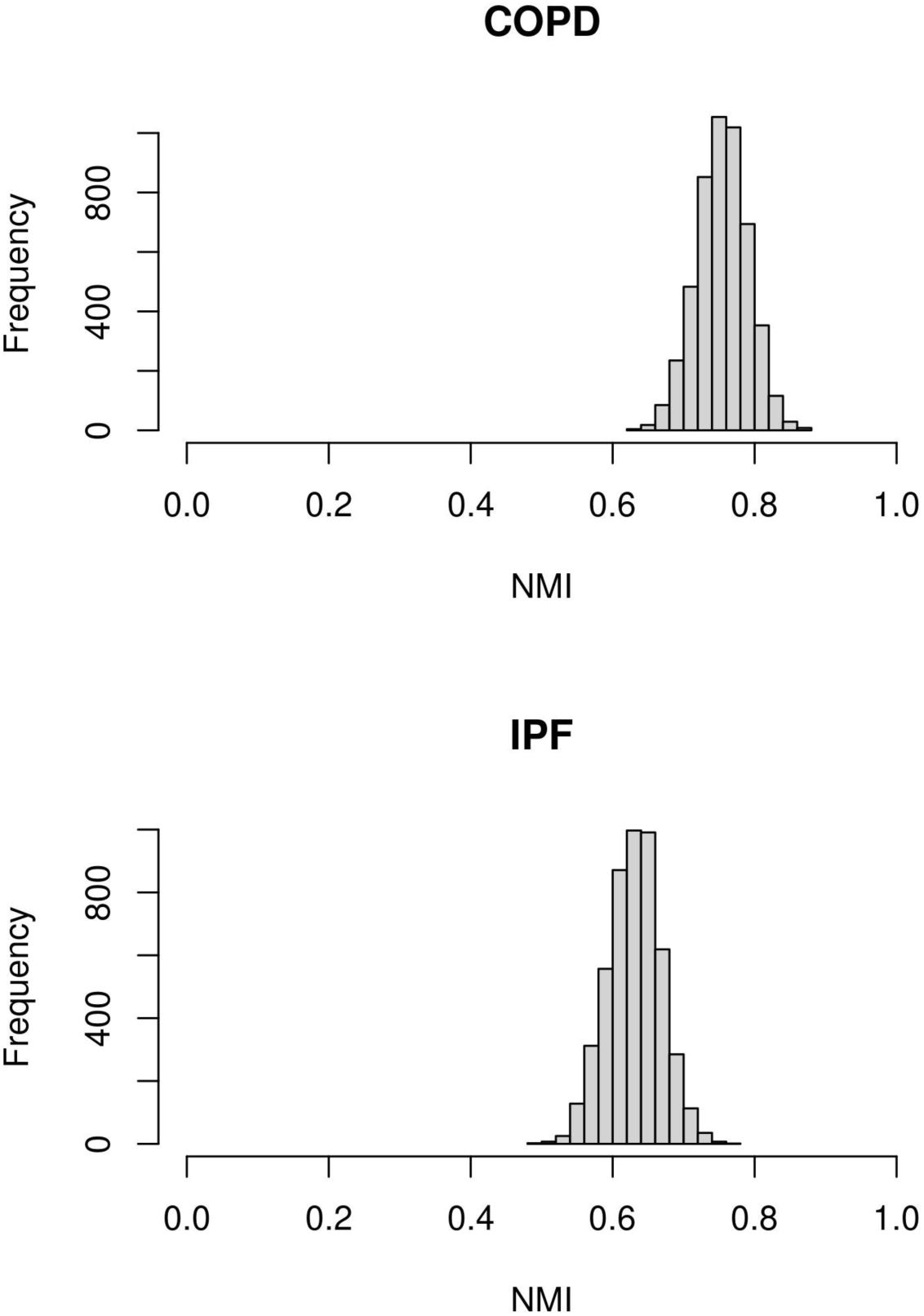
Normalized mutual information (NMI) between pairs of community structures estimated with Louvain clustering on the same dataset.

**Supplementary Figure 9:**
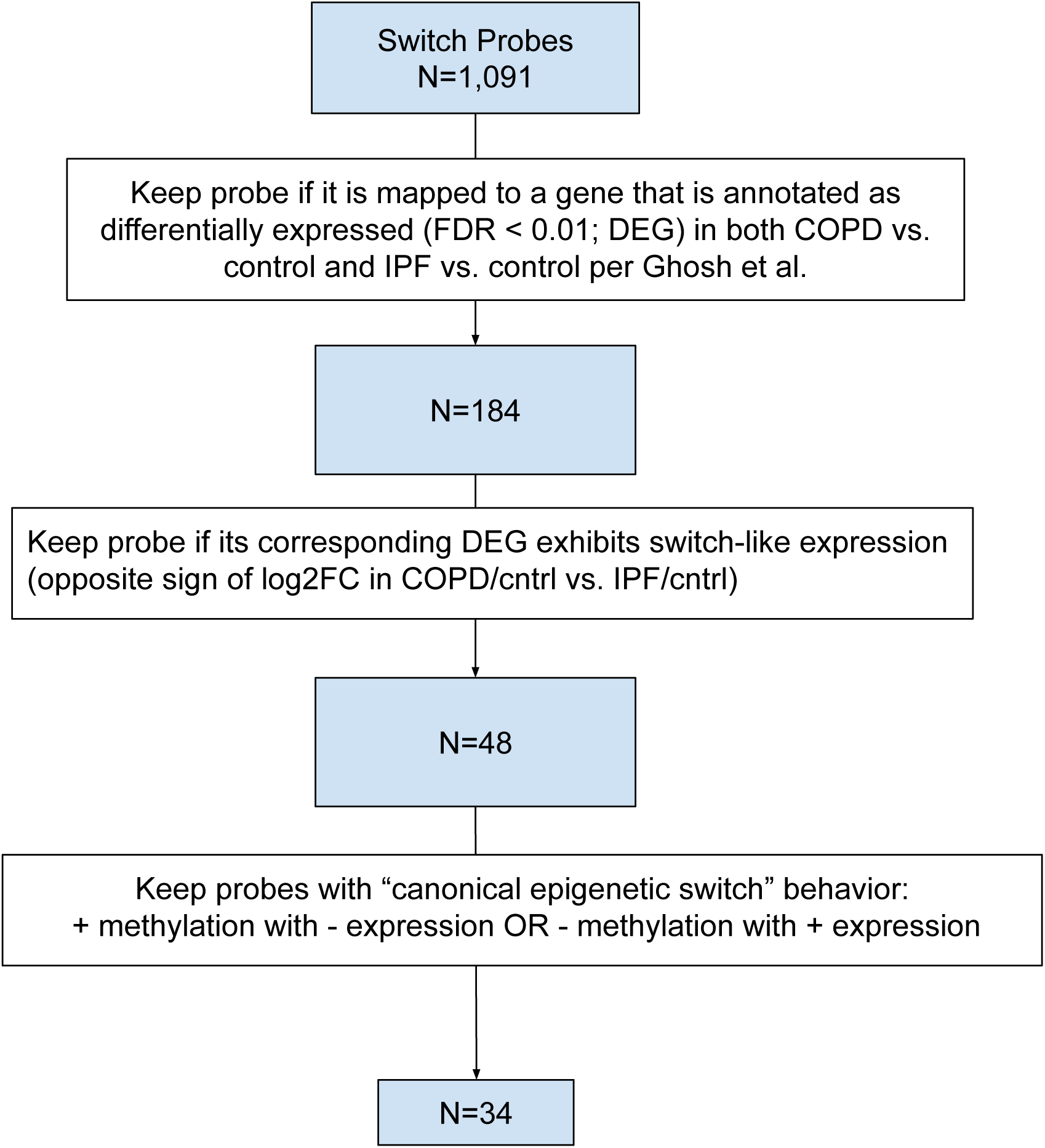
Flowchart for identifying canonical epigenetic switch probes (CES probes).

**Supplementary Figure 10:**
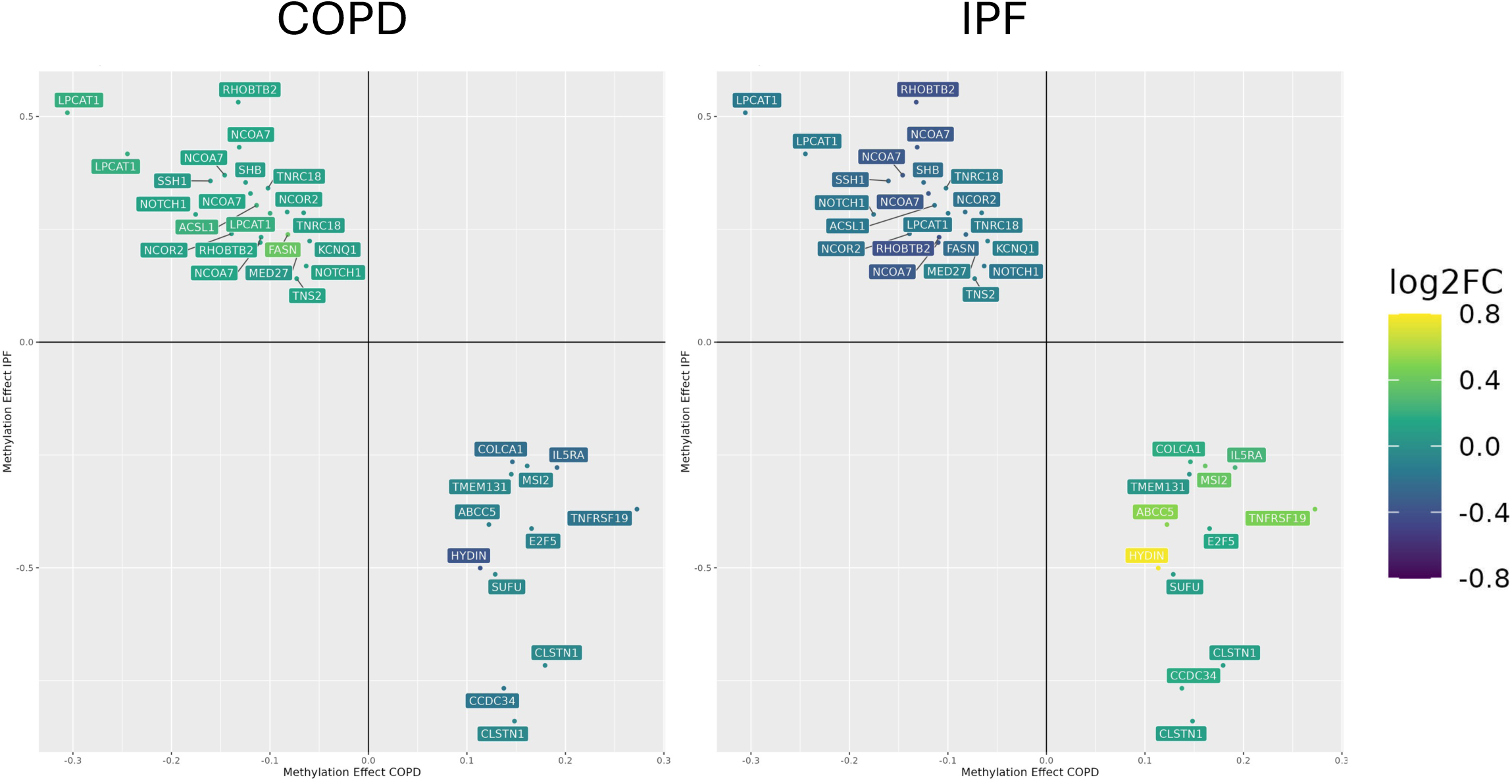
Gene expression of switch differentially expressed genes (DEGs) mapped to canonical epigenetic switch (CES) probes. Color scales and coordinate axes are the same on both plots.

